# Seroprevalence of SARS-CoV-2 antibodies prior to the widespread introduction of vaccine programmes in the WHO European Region, January - December 2020: a systematic review

**DOI:** 10.1101/2021.12.02.21266897

**Authors:** A Vaughan, EF Duffell, GS Friedl, DS Lemos, T Funk, A Nardone, M Valenciano, L Subissi, I Bergeri, E Broberg, P Penttinen, R Pebody, M Keramarou

**Affiliations:** World Health Organization Regional Office for Europe, Copenhagen, Denmark; European Centre for Disease Prevention and Control, Stockholm, Sweden; Epiconcept, Paris, France; World Health Organization, Geneva, Switzerland

## Abstract

**Background:** Seroprevalence surveys are essential to assess the age-specific prevalence of pre-existing cross-reactive antibodies in the population with the emergence of a novel pathogen; to measure population cumulative seroincidence of infection, and to contribute to estimating infection severity. With the emergence of SARS-CoV-2, ECDC and WHO Regional Office for Europe have supported Member States in undertaking standardized population-based SARS-CoV-2 seroprevalence surveys across the WHO European Region.

**Objectives:** The objective of this study was to undertake a systematic literature review of SARS-CoV-2 population seroprevalence studies undertaken in the WHO European Region to measure pre-existing and cumulative seropositivity prior to the roll out of vaccination programmes.

**Methods:** We systematically searched MEDLINE, ELSEVIER and the pre-print servers medRxiv and bioRxiv within the “COVID-19 Global literature on coronavirus disease” database using a predefined search strategy. We included seroepidemiology studies published before the widespread implementation of COVID-19 vaccination programmes in January 2021 among the general population and blood donors, at national and regional levels. Study risk of bias was assessed using a quality scoring system based on sample size, sampling and testing methodologies. Articles were supplemented with unpublished WHO-supported Unity-aligned seroprevalence studies and other studies reported directly to WHO Regional Office for Europe and ECDC.

**Results:** In total, 111 studies from 26 countries published or conducted between 01/01/2020 and 31/12/2020 across the WHO European Region were included. A significant heterogeneity in implementation was noted across the studies, with a paucity of studies from the east of the Region. Eighty-one (73%) studies were assessed to be of low to medium risk of bias. Overall, SARS-CoV-2 seropositivity prior to widespread community circulation was very low. National seroprevalence estimates after circulation started ranged from 0% to 51.3% (median 2.2% (IQR 0.7-5.2%); n=124), while sub-national estimates ranged from 0% to 52% (median 5.8% (IQR 2.3-12%); n=101), with the highest estimates in areas following widespread local transmission.

**Conclusions:** The review found evidence of low national SARS-CoV-2 seroprevalence (<10%) across the WHO European Region in 2020. The low levels of SARS-CoV-2 antibody in most populations prior to the start of vaccine programmes highlights the critical importance of vaccinating priority groups at risk of severe disease while maintaining reduced levels of transmission to minimize population morbidity and mortality.

## INTRODUCTION

The novel virus, Severe Acute Respiratory Syndrome–Coronavirus–2 (SARS-CoV-2) was first identified in Wuhan, China in December 2019 and spread rapidly around the world. At that time, the transmissibility, population susceptibility, clinical spectrum and infection-severity were all unknown. As of 5 November 2021, approximately 248 million confirmed cases and 5 million deaths have been reported globally, while in the WHO European Region, there have been 78 million cases and 1.4 million deaths (1, 2). However, notified cases and deaths are an underestimate of the true number of infections for reasons including clinical presentation with a large proportion of asymptomatic or mildly symptomatic cases, testing and reporting strategies and health care seeking behaviour (3). Asymptomatic infection has been reported in many studies with the proportion ranging from 6 to 41% (4-6), so a significant proportion of SARS-CoV-2 infections will be missed through case-based surveillance systems (7).

As the majority of infected individuals have a detectable humoral immune response on average 10-14 days after symptom onset and most individuals seroconvert within 3-4 weeks of infection (8), seroprevalence studies which measure SARS-CoV-2 antibodies can provide an important complement to routine surveillance, particularly as part of the assessment of novel emerging respiratory pathogens. Seroprevalence surveys are essential to assess the age-specific prevalence of pre-existing cross-reactive antibodies in the population; to measure population age-specific cumulative seroincidence as the novel virus spreads and to contribute to estimating infection-severity.

Since the start of the COVID-19 pandemic, there has been a rapid accumulation of seroepidemiological studies describing the seroprevalence of SARS-CoV-2. This review aims to provide a comprehensive review of studies conducted in the WHO European Region between 1 January and 31 December 2020 in the general population, with the aim to synthesize evidence on the extent of transmission across the region and population immunity to this newly emerging infection before the start of the COVID-19 vaccination programmes. As SARS-CoV-2 continues to circulate widely, understanding the age-specific population seropositivity remains critical for policymakers and public health officials to make informed decisions on optimal public health interventions (9).

## METHODS

### Search strategy

We searched MEDLINE, WHO COVID, ELSEVIER and the pre-print servers medRxiv and bioRxiv within the WHO “COVID-19 Global literature on coronavirus disease” database on 21/10/2020 and 12/01/2021. The searches spanned the period 1 January - 31 December 2020 and was not restricted by language. We supplemented these articles with WHO-supported Unity seroprevalence studies and unpublished studies reported to WHO Regional Office for Europe and ECDC. The selection process followed the Preferred Reporting Items for Systematic Reviews and Meta-analyses (PRISMA) guidelines (10). The full search strategy, search terms as well as inclusion and exclusion criteria are described in Supplementary Material S1.

### Data extraction

We combined the references from all databases, removed duplicates and imported the remaining articles into Rayyan software (11) for screening of titles and abstracts according to the inclusion and exclusion criteria (Supplementary Table S2). After the initial screening of title and abstracts, we assessed full-text publications for eligibility. At least two independent researchers extracted the eligible studies; a third researcher resolved any disagreements on assessment of eligibility or extraction. We extracted the following data: first author, publication date, country, region, period of study, population type, population age, sampling method, sample size, laboratory methods used, confirmatory testing, test performance, crude and adjusted point seroprevalence estimates, antibody type and analysis methodology. Comparison was made with weekly laboratory confirmed case and death reports. Data from pre-print articles and unpublished data included in this review were extracted and later updated if published article was available on 6 September 2021.

### Study quality assessment

We developed a quality assessment scoring system to assess the overall risk of bias of each study. The criteria included: a) the sampling frame (to assess representativeness of the general population); b) stratification (age, sex or population); c) recruitment method (random, convenience), d): adequacy of sample size, e): serological methods and validation; f) and statistical analyses (adjustment of results to account for the sensitivity and specificity of the test). A cumulative quality score classified the overall risk of bias of each study into high risk of bias (1-3), medium risk of bias (4-6) or low risk of bias (>6). See Supplementary Table S1 for more details on the quality criteria and Table 2 for scoring for each study. For the purposes of quality assessment, the threshold for acceptable test performance was ≥95% sensitivity and >97% specificity for laboratory assays and ≥90% sensitivity and >97% specificity for point-of-care tests (12).

### Data analysis

We used descriptive statistics to summarize results. We generated forest plots to display the data and explore variations according to specific characteristics, including time and population group. Correlation between cumulative incidence and cumulative deaths and seroprevalence estimates from studies of the general population was explored using Spearman’s rank correlation. We compared seroprevalence estimates from studies of the general population and the cumulative incidence and deaths at the start of each study. Analyses were performed in Microsoft Excel (version 2016) and R version 4.0.4.

## RESULTS

The literature search resulted in 4,063 studies. After deduplication, application of inclusion and exclusion criteria and supplementation with articles from other sources, a total of 111 studies were included in this review. Of these, 77 were published articles, 19 were preprints, nine were institutional reports, and six were studies were identified through reporting of unpublished results to WHO or ECDC. See Figure 1 PRISMA flow diagram study selection.

**Figure 1:**
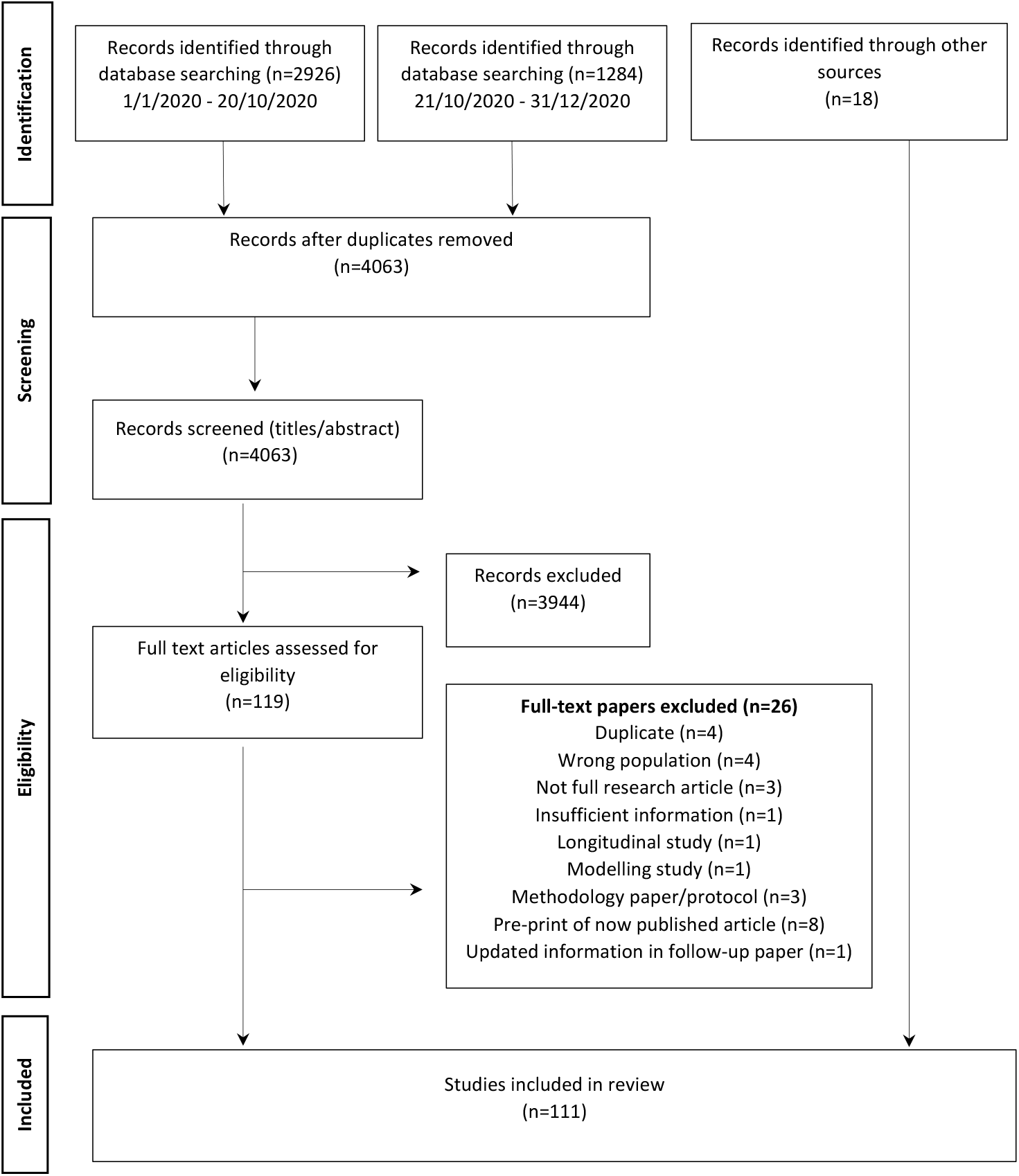
PRISMA Flow chart of SARS-CoV-2 seroprevalence study selection.

The 111 studies included 224 seroprevalence estimates from 26 of the 53 countries in the WHO European Region (Figure 2). The majority of studies (n=82; 74%) were conducted in 19 EU/EEA countries, while 29 studies (26%) conducted in seven non-EU/EEA countries (Bosnia and Herzegovina, Georgia, Kyrgyzstan, Republic of Moldova, Russian Federation, Switzerland and the United Kingdom) (Figure 2; Table 1). Fifty-six (50%) studies were aligned with the World Health Organization Unity population-based sero-epidemiological investigation criteria related to study design, data collection and analysis (2).

**Table 1:** Study characteristics.

| Characteristics | Number of studies | % |
| --- | --- | --- |
| **Total** | 111 | 100 |
| **Study characteristics** |  |  |
| **Country** |  |  |
| WHO European Region (EU/EEA) | 82 | 74 |
| WHO European Region (outside of EU/EEA) | 29 | 26 |
| **WHO UNITY alignment** |  |  |
| Unity-aligned | 56 | 50 |
| Not Unity-aligned | 55 | 50 |
| **Publication type** |  |  |
| Peer-reviewed article | 77 | 69 |
| Pre-print | 19 | 17 |
| Institutional report | 9 | 8 |
| Not yet published | 6 | 5 |
| **Geographical level** |  |  |
| National | 33 | 30 |
| Regional | 27 | 24 |
| City/Local | 50 | 44 |
| Multiple | 1 | 1 |
| **Sampling strategy** |  |  |
| Convenience | 69 | 62 |
| Random | 41 | 37 |
| Not reported | 1 | 1 |
| **Population type** |  |  |
| Household/Community only | 45 | 41 |
| Residual sera only | 13 | 12 |
| Blood donors only | 16 | 14 |
| Patients seeking healthcare (non COVID-19) only | 13 | 12 |
| Pregnant or parturient women only | 7 | 6 |
| Other/multiple | 23 | 21 |
| **Quality assessment** |  |  |
| Low risk of bias | 41 | 37 |
| Medium risk of bias | 40 | 36 |
| High risk of bias | 24 | 22 |
| N/A | 6 | 5 |
| **Sample size** |  |  |
| <1000 | 45 | 41 |
| >=1000 | 66 | 59 |
| **Laboratory characteristics** |  |  |
| **Serological method** |  |  |
| ELISA | 55 | 50 |
| CMIA/CLIA | 42 | 38 |
| LFA | 25 | 23 |
| MN | 10 | 9 |
| Other | 8 | 7 |
| Not reported | 2 | 1 |
| **Type of assay** |  |  |
| Commercial | 90 | 81 |
| In-house | 26 | 23 |
| Not reported | 2 | 1 |

\*ELISA – Enzyme linked immunoassay; CMIA/CLIA - Chemiluminescence Microparticle Immunoassay/ Chemiluminescence Microparticle Immunoassay; LFA – Lateral flow immunoassay; MN – Microneutralization assay

**Table 2.**
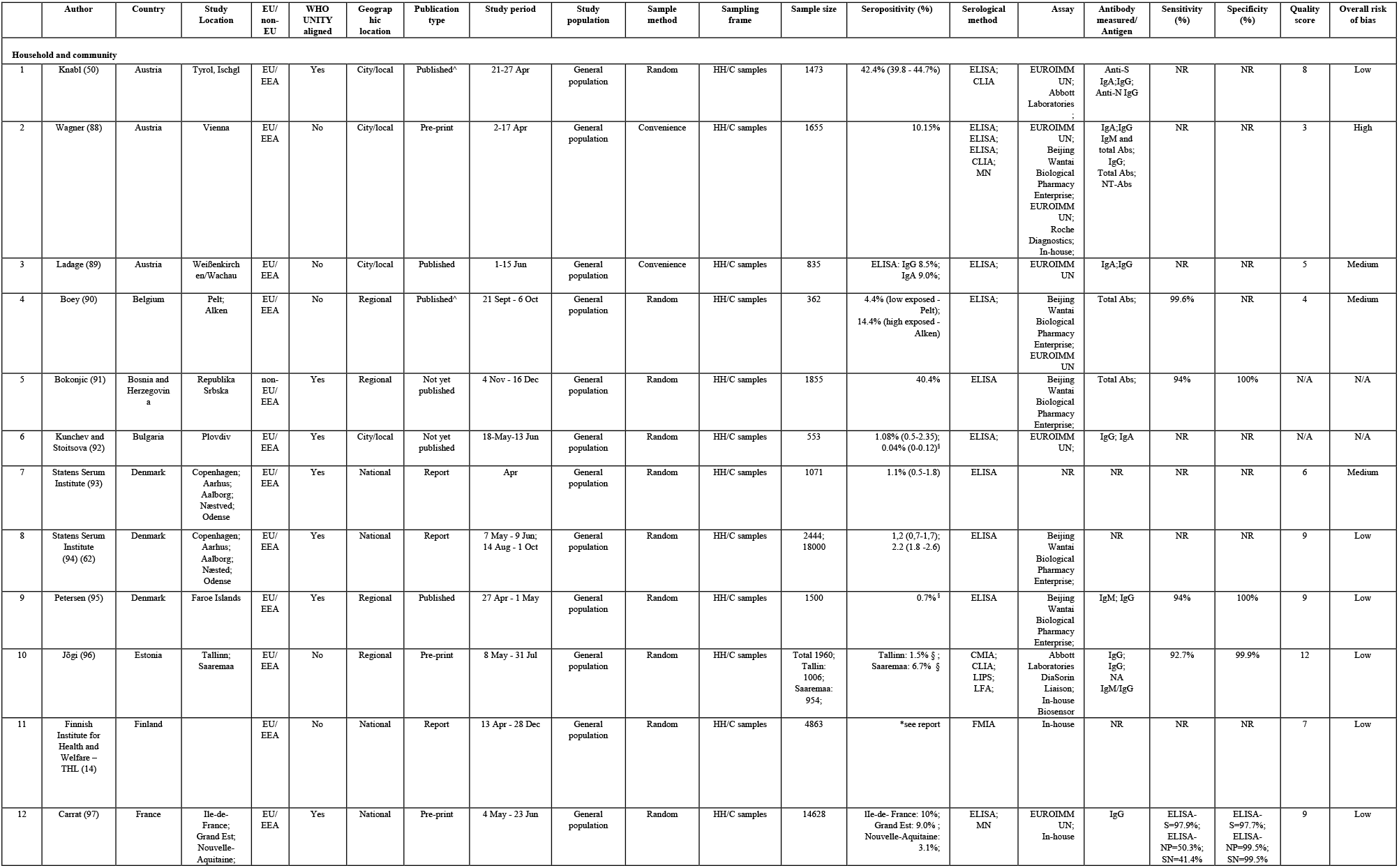
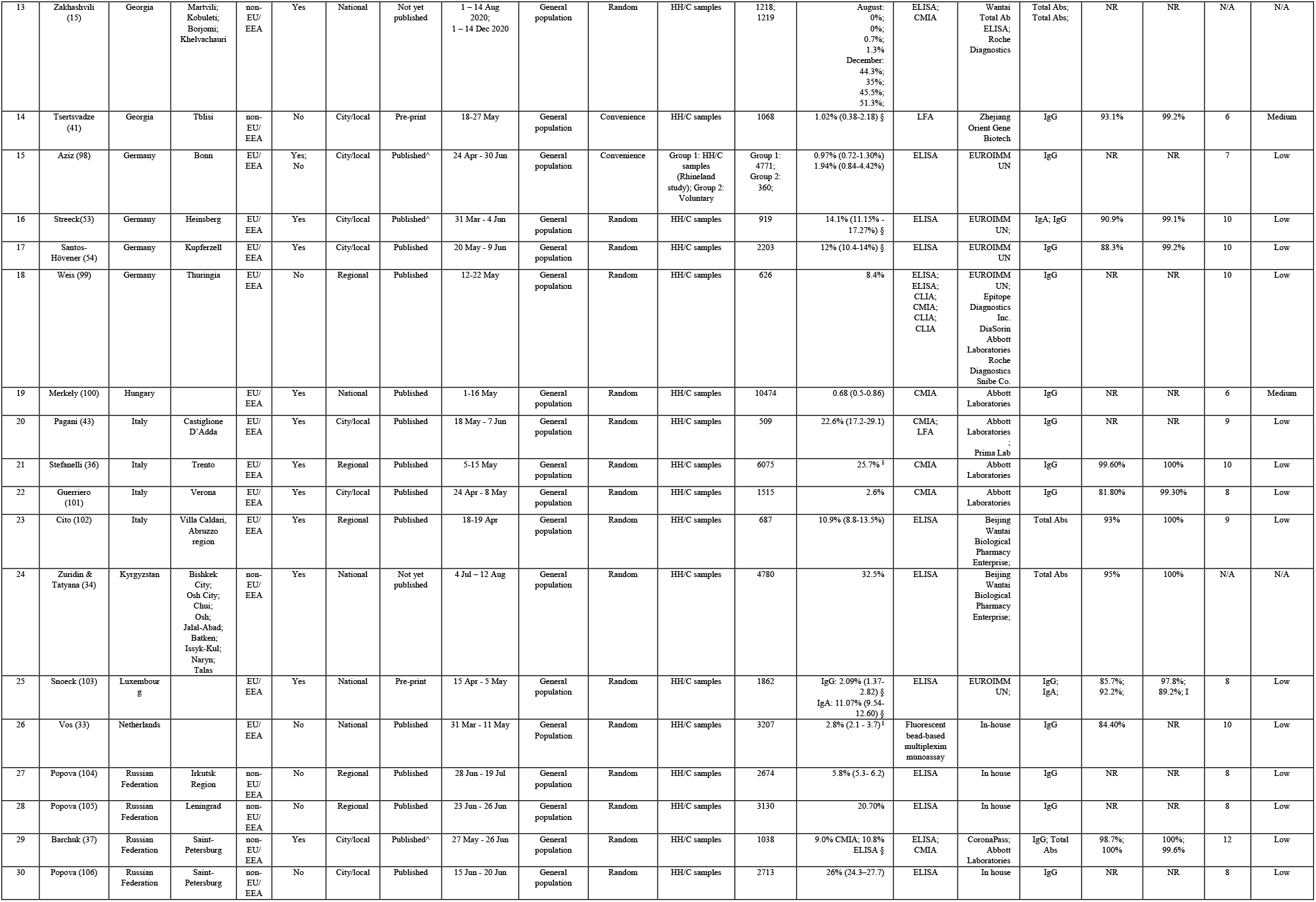
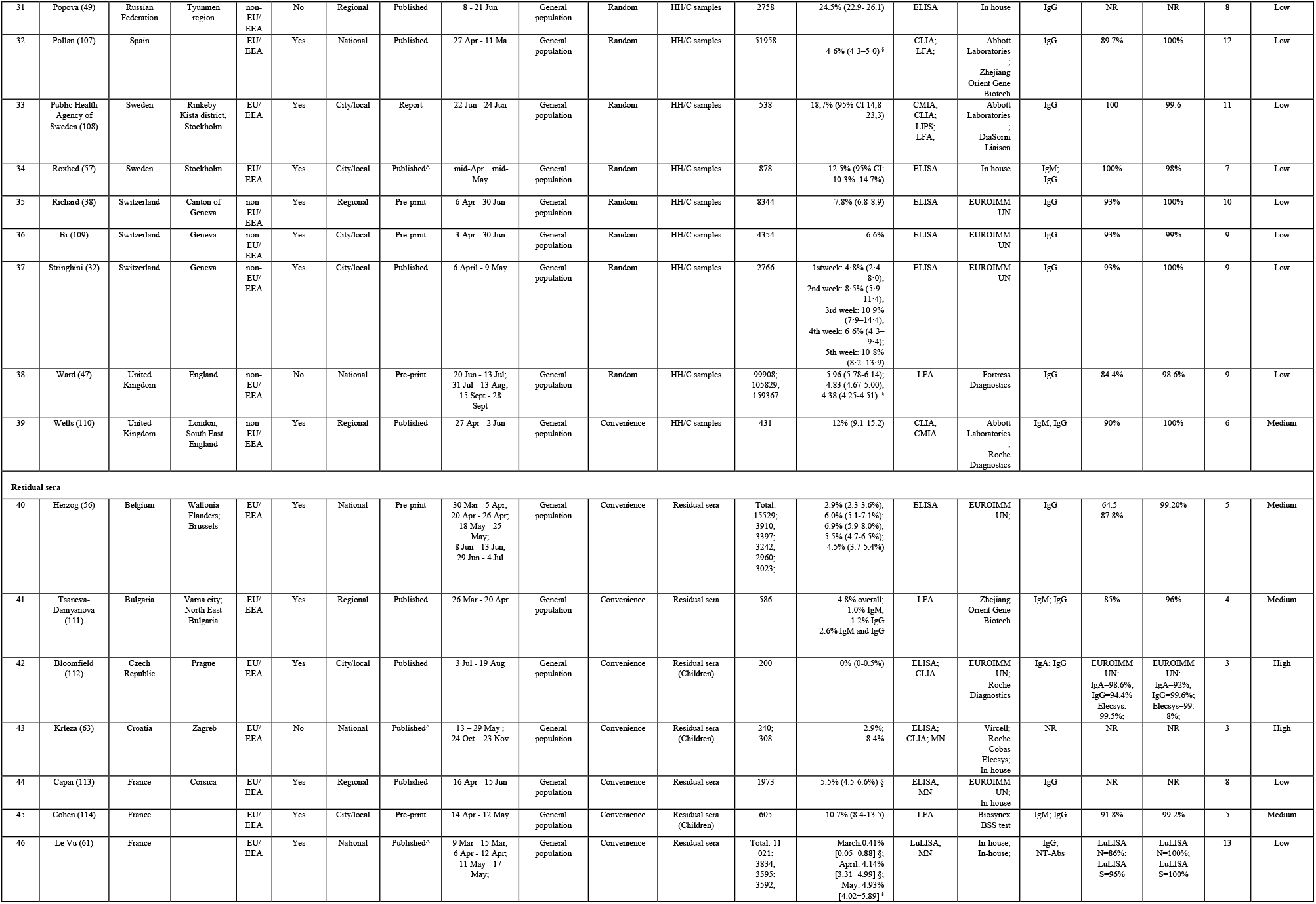
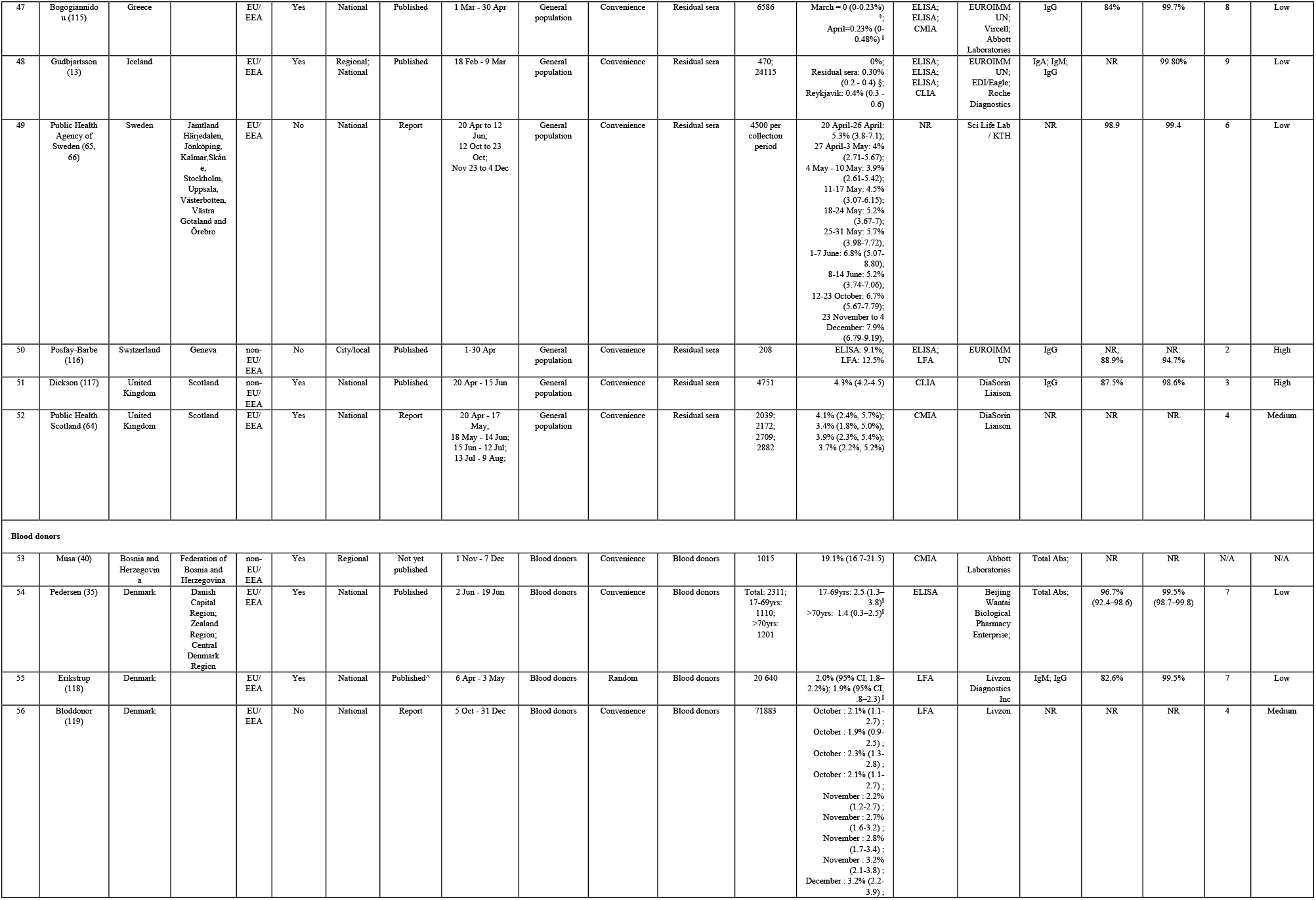
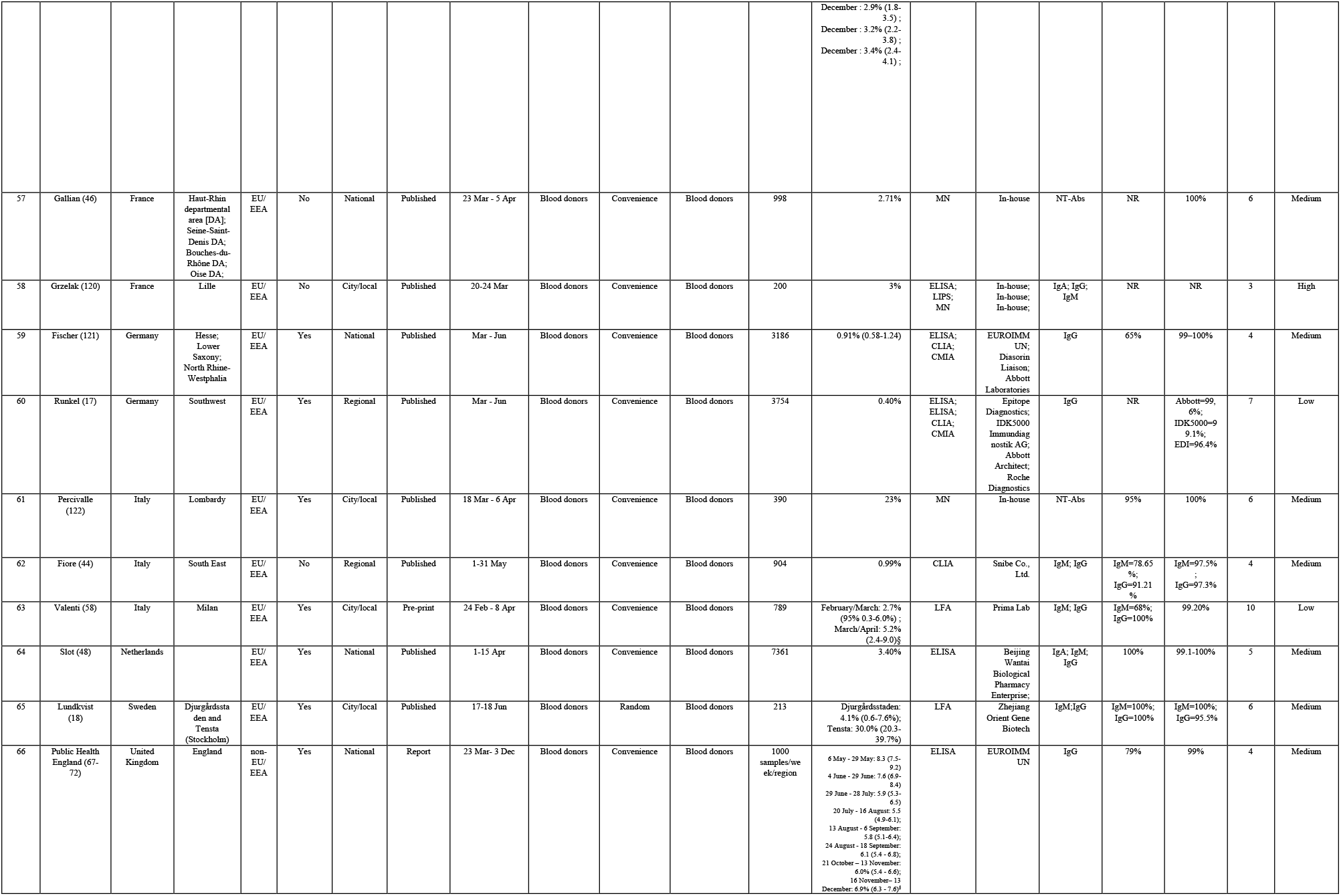
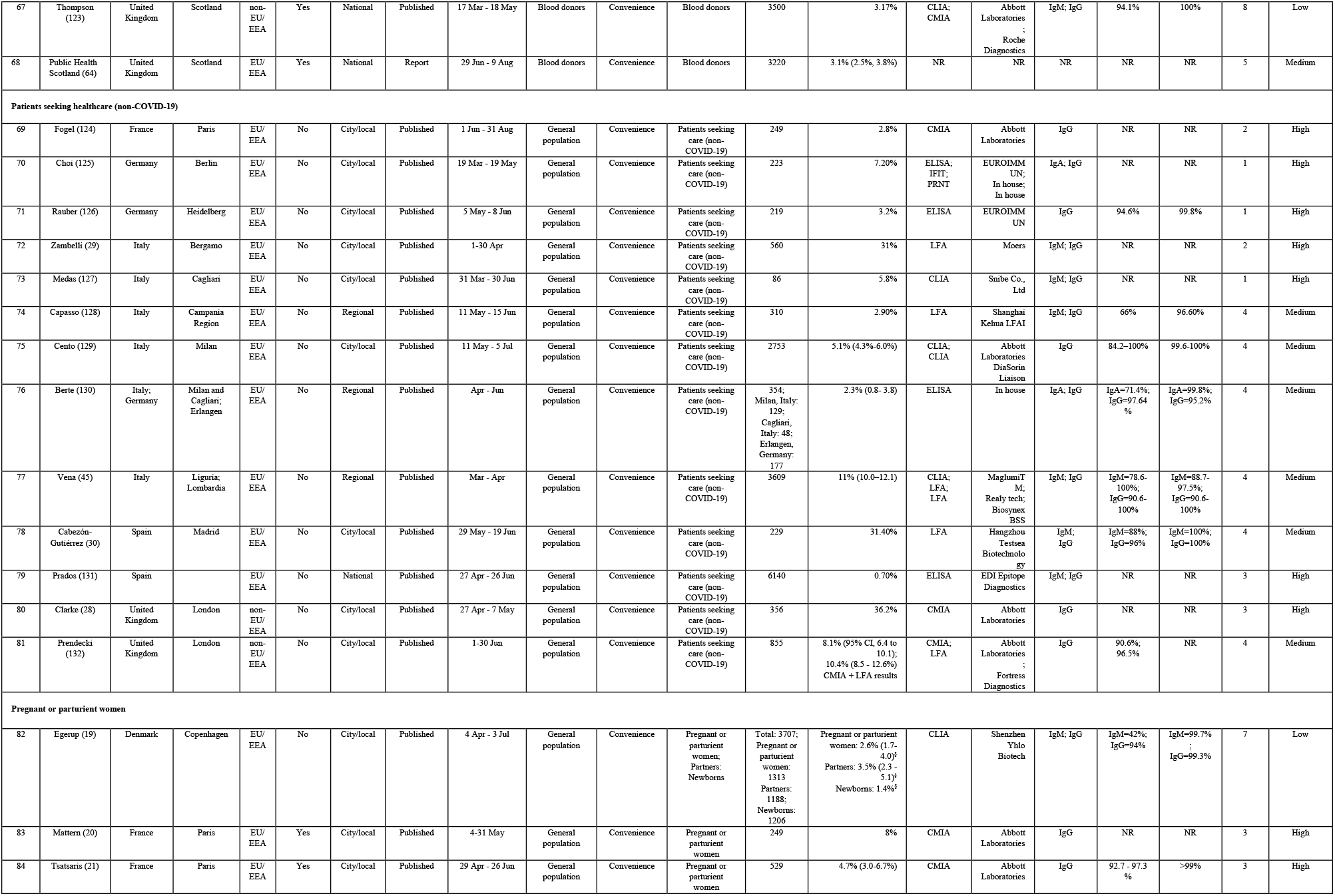
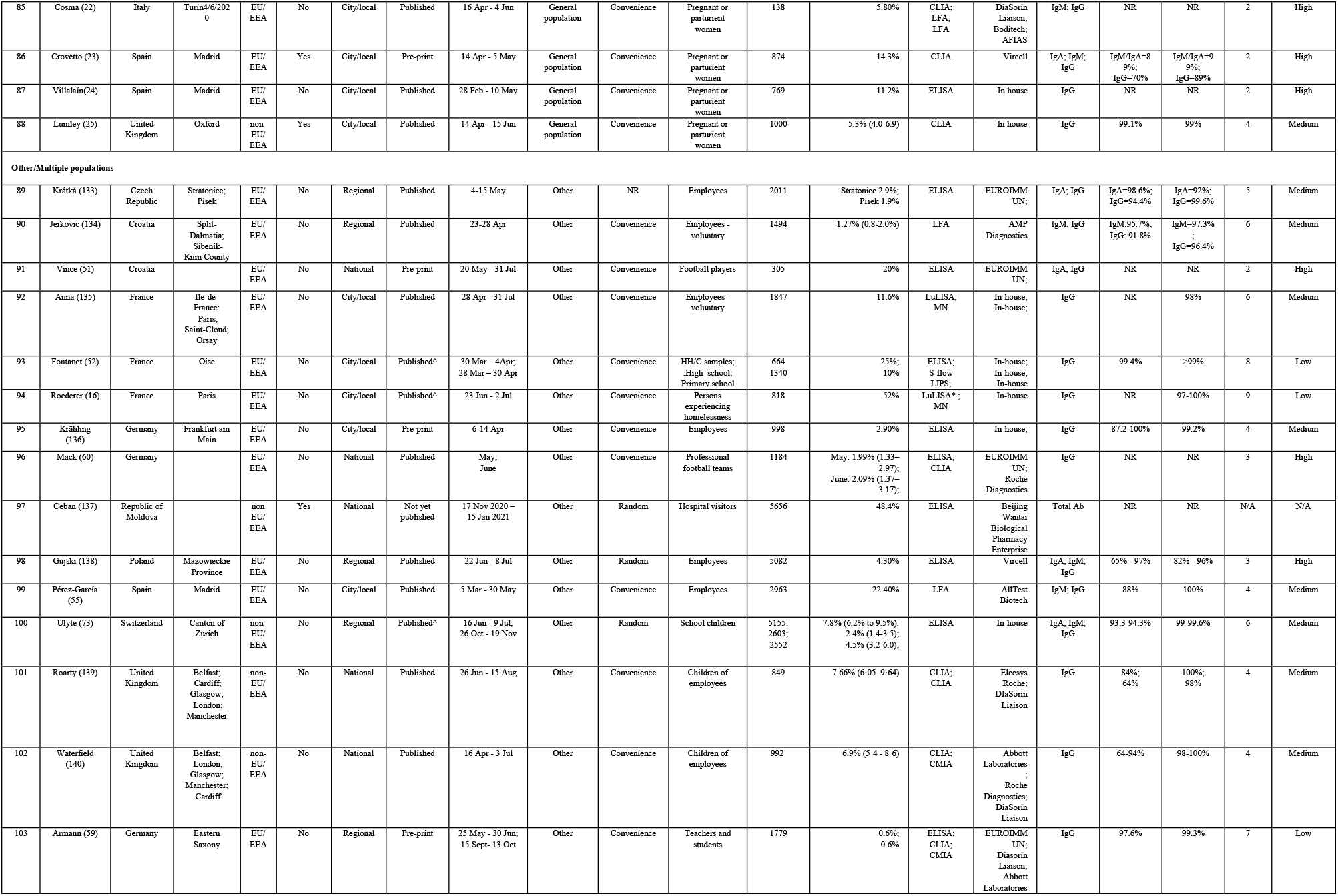
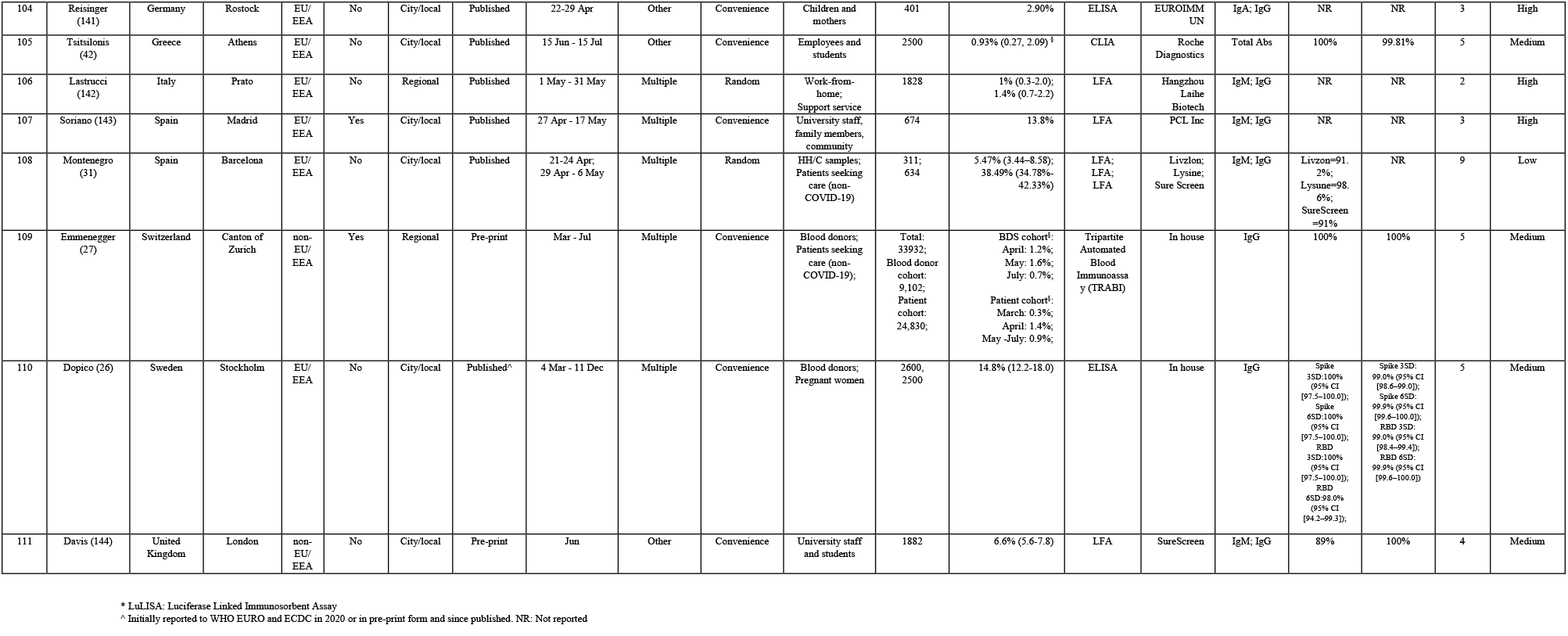
Characteristics of eligible seroprevalence studies.

**Figure 2:**
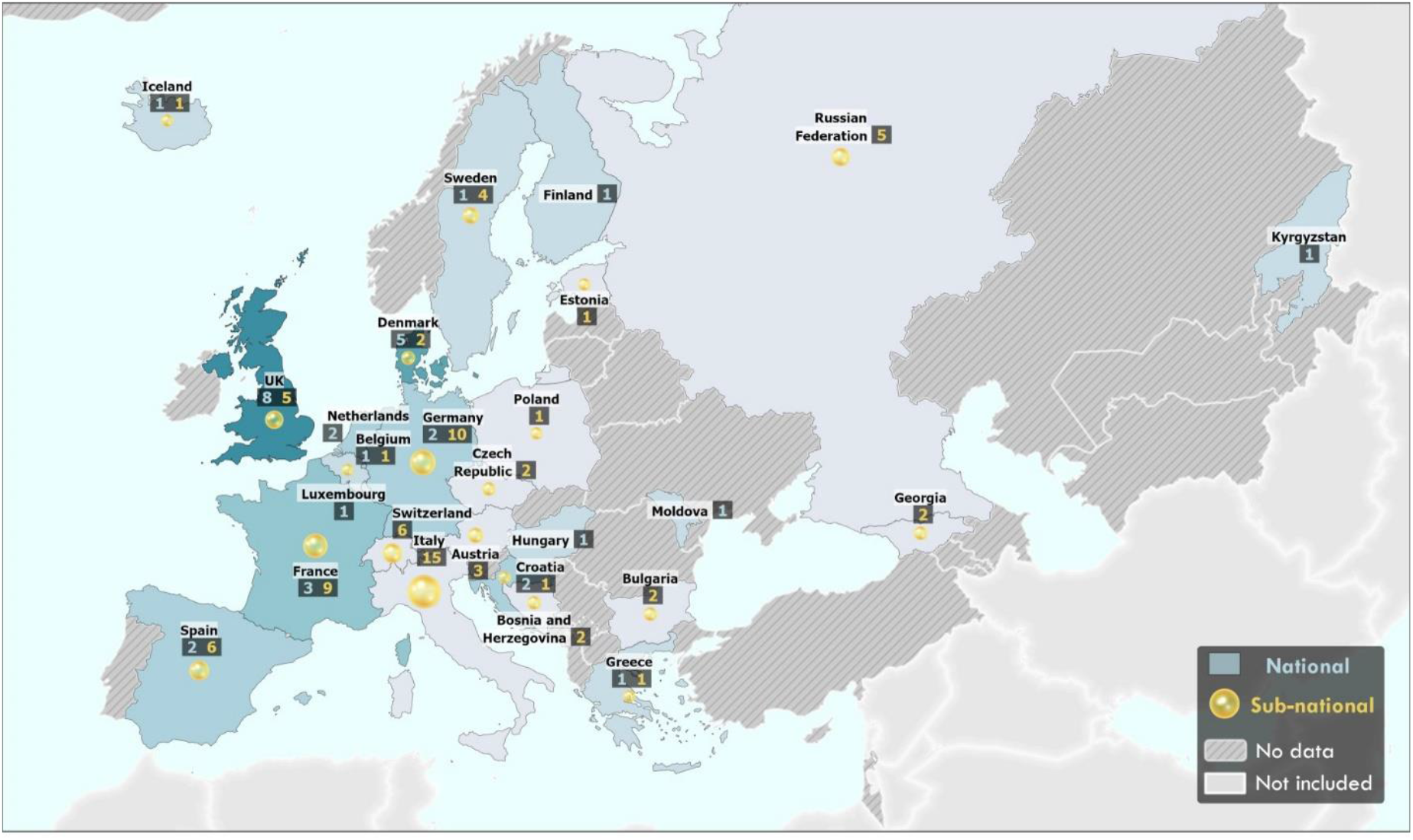
Geographical distribution of SARS-CoV-2 seroprevalence studies published in the WHO European Region between 1 Jan – 31 Dec 2020. Countries with national-level seroprevalence studies are reported in blue (shade of blue reflects the number of studies conducted in the country/territory). Subnational-level seroprevalence studies are reported as a yellow circle (Size of circle reflects number of subnational studies conducted in the country/territory). Number of studies are listed in boxes under name. Countries with not studies are coloured in grey. The designations employed and the presentation of this material do not imply the expression of any opinion whatsoever on the part of the Secretariat of the World Health Organization concerning the legal status of any country, territory, city or area or of its authorities, or concerning the delimitation of its frontiers and boundaries. Dotted and dashed lines on maps represent approximate locations for which there may not yet be full agreement.

The majority of studies (n=69, 62%) used non-random or convenience sampling of the population. Forty-one (37%) studies used random sampling, while one study did not report sampling methodology. Characteristics and details of included studies are shown in Table 1 and Table 4, respectively.

In total, 72 (65%) of the studies provided representative estimates from the general population, of which sample frames included 45 (41%) studies of household or community samples, 13 (12%) residual sera, 13 (12%) patients seeking healthcare for non-COVID-19 related issues, seven (6%) pregnant or parturient women. Sixteen (14%) studies sampled blood donors as a proxy for the general population while 23 (21%) sampled other or multiple populations. Studies were conducted at differing geographical levels within a country, including at the national level (n=33; 30%), regional level (n=27; 24%) and city or local level (n=50; 44%). One study reported both national and regional estimates (13).

Over half of the studies used one serological assay (74; 67%) while 34 (31%) used at least two different assays. In 82 studies (74%), commercial assays from various sources were used, 20 (18%) studies used an in-house assay only and six studies (5%) used both a commercial and in-house developed assay. The test method was not reported in two studies. An Enzyme-linked Immunosorbent assay (ELISA) was the method most commonly employed (n=55, 50%), followed by Chemiluminescent immunoassay (CLIA) or Chemiluminescence Microparticle Immunoassay (CMIA) (n=42, 38%) and lateral flow immunoassays (LFAs) (n=25, 23%). Seventeen studies (15%) used LFAs exclusively. Ten studies (9%) employed in-house microneutralization assays to assess the neutralizing ability of SARS-CoV-2 antibodies.

Of 90 studies that used a commercial assay, 33 studies (37%) reported the use of tests with acceptable sensitivity and specificity. Of those that independently validated assay performance (n=41, 46%), 14 (34%) reported acceptable sensitivity and specificity, while 27 (66%) did not meet these thresholds. Of the 20 studies that used an in-house assay, nine (45%) reported an acceptable test performance, four (20%) performed below these thresholds and seven (35%) did not report on test performance. The majority of studies (n=83, 75%) did not report adjustment for test sensitivity or specificity in their analysis.

Based on our quality scoring system (Supplementary Table S3), 81 studies (73%) were of high or medium quality reflecting a low or medium risk of bias, respectively (medium quality: n=40, 36%; high quality n=41, 37%). A total of 24 studies (22%) were determined to be at high risk of bias, largely due to non-random sampling frame, weak representativeness of the general population or lack of adjustment for sampling bias or test performance. A quality assessment was not performed for unpublished studies.

Seroprevalence estimates (n=88) from national studies ranged from 0% (CI: 0.0-0.7) in Finland in May (14) to 51.3% in Georgia in December (15) (median 2.2% (IQR 0.7 – 5.2%); n=124) (Figure 3a), while seroprevalence estimates from studies spanning regions, cities or towns (n=101) ranged from 0% (CI 0.0-0.5%) in Czech Republic in August 2020 (15) to 52% in a Médecins Sans Frontières centre in Paris, France during an outbreak with widespread community transmission in June 2020 (16) (median 5.8% (IQR 2.3-12%); n=101) (Figure 3b).

**Figure 3:**
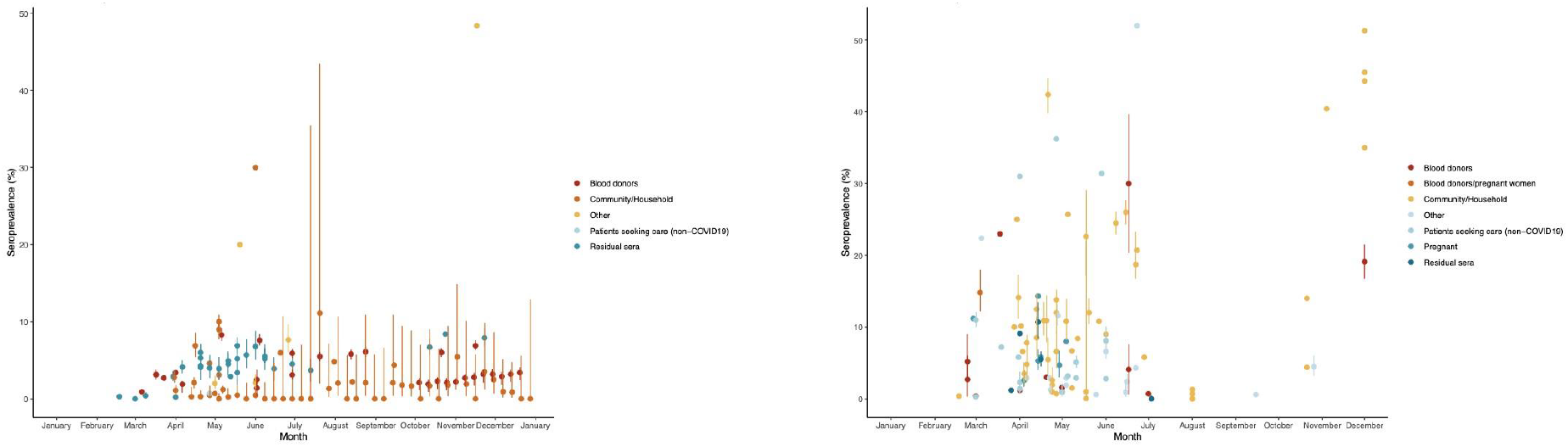
National (a) and sub-national (b) seroprevalence estimates of SARS-CoV-2 antibodies over time in the WHO European Region (1/1/2020-31/12/2021)

A total of 45 studies provided seroprevalence estimates (n=105) from community or household samples and 39 studies (87%) were found to be of high or medium quality. Seroprevalence estimates ranged from 0% (CI: 0-0.7%) in Finland in May and to 51.3% in December 2020 in Georgia (15) (median 2.6% (IQR 0.5-10%) n=105) (Supplementary Figure S1).

Thirteen studies screened residual clinical samples (29-42) between February and November 2020, of which nine (70%) were of high or medium quality. Seroprevalence estimates (n=34) in this population varied across countries ranging from 0% (CI 0-0.23) in Greece in March to 18.7% (CI 16.7-23.3%) in Sweden in June (median 4.5% (IQR 3.5-5.9%); n=34) (Supplementary Figure S2a).

Eighteen studies (17%) utilized blood donors as a proxy for the general population between February and December 2020, of which 16 were of high or medium quality. Seroprevalence estimates (n=42) in blood donors varied across countries, ranging from 0.4% in Germany between March and June (17) to 30% in Tensta (Stockholm) following a period of high incidence in June (18) (median 5.8% (IQR 2.1-5.7%) n=42) (Supplementary Figure S2b).

Eight studies investigated the seroprevalence of SARS-CoV-2 in pregnant or parturient women, reporting estimates ranging from 2.6% (CI 1.7-4%) and 14.3% between March and June 2020 (median 6.9% (IQR 5.1-12%); n=8) (19-25) (Supplementary Figure S2c). One study provided combined estimates of blood donors and pregnant women of 14.8% in Sweden between March and December (26) Fourteen studies provided 16 estimates from individuals seeking healthcare for non-COVID-19 related reasons and seven (50%) of these were medium or high quality. Estimates ranged from 0.3% in Zurich, Switzerland in March (27) to 36.2% in London in April (28) (median 4.1% (IQR 2.1-8.8%); n=16) from March to August 2020. The highest seroprevalence estimates (>10%) in this group were observed in three patient groups investigated following local widespread community transmission, oncology patients (31%) in Bergamo, Italy in April 2020 (29), oncology patients (31.4%) in Madrid between May and June 2020 (30) and haemodialysis patients (36.2%) in London in April and May 2020 (28) and patients (38.5%) in Barcelona, Spain in April (31) (Supplementary Figure S2d).

Forty-four (41%) studies reported seroprevalence estimates stratified by age. Seroprevalence estimates varied considerably across age groups and estimates tended to be lower in children (<18 years) (32-34) and older age groups (>60 years) (32, 35-40). Whilst a number of studies reported a high seroprevalence in older age groups (>55 years) (36, 37, 41-46), some studies also reported a higher seroprevalence in younger age groups (<40 years) (33, 46-48). In studies that reported seroprevalence estimates by sex, similar seroprevalence results were observed between females and males with the exception of a study in Italy (45), Russian Federation (49) and Kyrgyzstan (34) which each found a higher seroprevalence in females.

A number of studies provided seroprevalence estimates prior to, or at the early stages of the epidemic in the country (Supplementary Figure S3). Of these, overall study estimates were largely below 10%, however higher seroprevalence was noted in a number of population-specific, regional or local studies (29, 36, 43, 50-54), with suggestion of earlier undetected transmission in some countries (24, 26, 34, 55). A total of 16 studies reported seroprevalence estimates spanning multiple timepoints or stages of the epidemic (14, 15, 27, 31, 32, 47, 56-73). In a serial cross-sectional study in France (61), residual blood sampled before, during and after a national lockdown showed a seroprevalence of 0.41%, 4.14%, and 4.93%, respectively. In Georgia, in a community sample, an increase in seroprevalence from 0-1.3% in August 2020 to 35-51.3% in the same regions in December 2020 was noted (15). A seroprevalence study in blood donors conducted in Milan between February and April 2020 during a period of intense transmission found an increase in seroprevalence from 2.7% (95% 0.3-6.0%) to 5.2% (95% 2.4-9.0), with an adjusted rate of increase in antibodies (IgG) of 2.7±1.3% per week as social distancing measures were gradually implemented (58). While in Finland, weekly testing of blood donors from April 2020 onwards showed a consistently low seroprevalence in the general population over time (0.28% (0.05– 1.55) in early April 2020 to 0% (0–12.87) in late December 2020 (67).

The relationship between seroprevalence and reported SARS-CoV-2 laboratory confirmed cumulative case and deaths incidence was also explored. While seroprevalence from national studies correlated moderately with cumulative incidence (Spearman’s rank correlation coefficient, 0·52) (Figure 4a), a stronger correlation was observed between seroprevalence estimates and cumulative SARS-CoV-2 deaths (Spearman’s rank correlation coefficient, 0·754) (Figure 4b).

**Figure 4:**
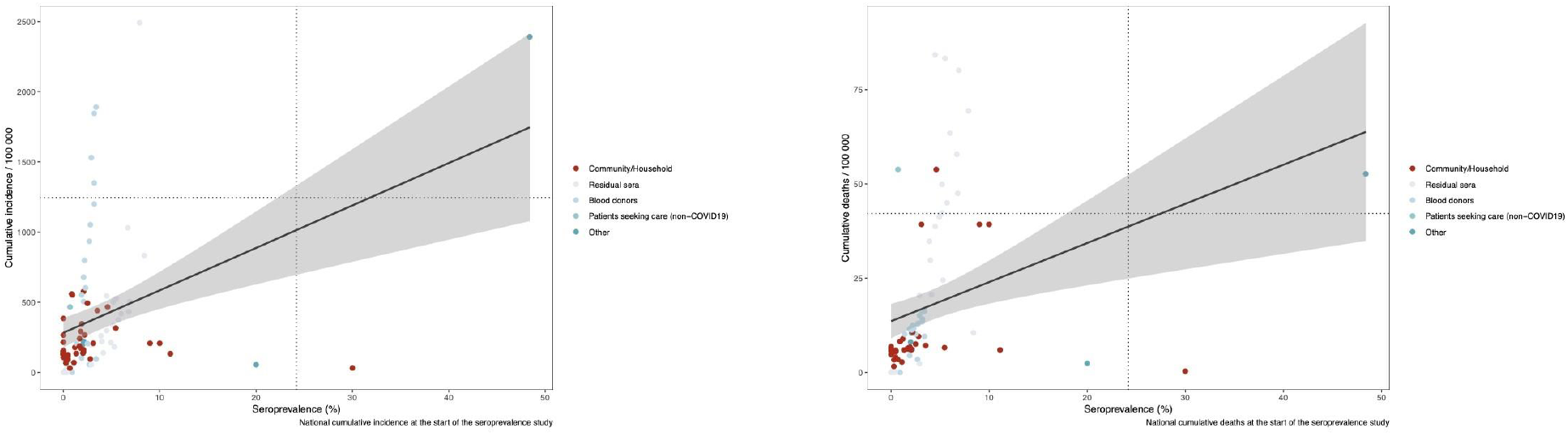
Correlation between seroprevalence point estimates from low to medium risk of bias studies and cumulative (a) incidence and (b) deaths in all populations, in the WHO European Region (1/1/2020-31/12/2020)

## DISCUSSION

In this study we report the results of 111 studies, including 224 seroprevalence estimates from 26 countries in the WHO European Region undertaken until December 2020, prior to the implementation of national COVID-19 vaccine campaigns. A significant heterogeneity in implementation was noted across the studies, with a bias towards studies in high-income countries in Western Europe.

Overall, population-wide seroprevalence estimates were low (below 10%) across the Region early in 2020 before the onset of widespread community transmission and remained low across the Region throughout 2020, despite circulation of SARS-CoV-2 over this period. Higher estimates were observed at a regional or local level in populations that had experienced intense community transmission (up to 52%). Furthermore, a positive correlation between seroprevalence estimates and national cumulative incidence was observed, with a stronger correlation between seroprevalence and cumulative mortality.

The wide variation in seroprevalence estimates across the region are likely to reflect many factors including the differences in the population studied, local stage of the epidemic and the public health and social measures implemented in response to the epidemic at that time. The general low seroprevalence both at the start of the pandemic and at the end of 2020 is in line with a number of global systematic review conducted to date (74-76) and together indicate that the majority of the proportion of the population in the WHO European Region were and remain susceptible to infection one year after the identification of SARS-CoV-2 and prior to the start of national vaccination campaigns. In a global systematic review, Chen et al. estimated a seroprevalence of 4.2% (2.7-5.8) across the European Region until August 2020 (76) while Rostami et al. estimated a pooled prevalence of 3.17% (1.96-4.38), 4.41 % (2.20-6.61), 5.27% (3.97-6.57) in Western, Southern and Northern Europe, respectively (75). In the same period, Bobrovitz et al. reported a pooled estimate of 1.6% (1.1-5.2%) seroprevalence in studies conducted across Central Europe, Eastern Europe and Central Asia (77) and 12.2% (4.5-25.4%) from population-wide studies conducted until December 2020 (74).

A number of studies reported low seroprevalence in younger and older age groups, a finding observed in other systematic reviews (74, 76, 78). Such findings have important implications, as groups such as the elderly are at higher risk of severe outcome following infection – and lack of cross-protective immunity indicates that all age-groups will anticipate to see high infection attack rates without implementation of measures such as vaccination of priority groups, together with strengthening of public health and social measures to reduce SARS-CoV-2 transmission.

When reviewed alongside case notification data, seroprevalence estimates can provide greater insight into the local evolution of the pandemic. In this review, a positive correlation between seroprevalence estimates and national cumulative incidence in a number of countries was observed, suggesting that seroprevalence is a reflection of the duration and intensity of community transmission. It should be noted however that during the initial peak of infections in Europe in the spring of 2020, testing in many countries was not yet optimal and case notification data at this time are unlikely to provide a robust proxy for incidence in many instances. In line with this, several studies found seroprevalence estimates to be higher than the corresponding cumulative incidence of SARS-CoV-2 infections, suggesting a substantial under-ascertainment of infection through notifications, due to a number of factors including the asymptomatic or mild nature of disease, healthcare seeking behaviour, lack of testing capacity and testing and reporting strategies. Indeed, we also found a stronger association between seroprevalence and cumulative case mortality than cumulative case incidence, providing further evidence to support the suggestion of case under-ascertainment, as laboratory confirmed mortality surveillance for COVID-19 is likely to be more comprehensive.

The varying quality of studies in this review reflects the challenge of conducting seroepidemiological studies of high quality. Indeed, this review found that only 50% of all studies undertaken in the WHO European region in 2020 were aligned with the WHO Unity study initiative. Few of the national (n=5; 15%) or regional (n=2; 7%) studies were determined to be of high risk of bias, while 17 (34%) of studies conducted at a local level (cities or towns) were graded as such. This variation may be explained by the level of resources and epidemiological support available to studies conducted at the regional or national level.

The majority of studies identified in this review utilised convenience rather than random sampling, which may have reduced the true representativeness of the estimates derived, though such convenience sampling is likely to provide a good estimate of population exposure for widely circulating viral infections. Many studies also included individuals that were not fully representative of the population under study, which may have introduced bias. For example, this review included studies that explored seroprevalence in the general population by utilising various proxy populations such as blood donors and residual blood. Blood donors are known to differ from the general population in that they are often a young, healthy adult population selected on the basis of lack of recent infection (79) and seroprevalence may therefore be over or underestimated in this group. Residual sera, on the other hand, derives from individuals who have sought health care and may therefore have pre-existing comorbidities or be at higher risk of SARS-CoV-2 infection. However, we found that seroprevalence estimates for these distinct populations are in good agreement with the general population.

We also found that there was a high degree of heterogeneity across serological assays used. The majority of studies used commercial tests of varying sensitivity and specificity to detect SARS-CoV-2 targeted antibodies, although some of these assays have now been shown to have excellent performance (80, 81). However, under half of studies performed independent validation of these kits with internal controls and serum panels and only 25% accounted for the sensitivity and specificity of the tests in their statistical analyses. As SARS-CoV-2 serological tests have been found to have variable test performance (80, 81), independent validation at local level in combination with use of an WHO International Standard and Reference Panel for anti-SARS-CoV-2 antibody has been widely promoted as part of the Solidarity II initiative (82, 83). Other options include the Joint Research Centre (84) reference materials for the quality control of SARS-CoV-2 antibody tests. Use of these materials will allow for the potential correction for sensitivity and specificity during the statistical analysis, would allow for more robust estimates and greater comparability among countries in the region.

Overall, the findings of this review highlight the need for international collaboration to standardise approaches and support countries in conducting robust comparable studies. WHO, in collaboration with technical partners, has developed the Unity studies (2)(90), a global seroepidemiology standardization initiative for COVID-19, which aims to increase quality evidence-based knowledge in country and regions for action through the availability of standardized seroepidemiology investigation protocols and antibody assays. A primary aim of this global initiative is the provision of direct support to countries to develop country specific protocols, with particular attention provided to low- and middle-income countries (LMICs), and to support aggregation, comparison and analysis of robust Unity-aligned studies through strong coordination between WHO Country offices, Regional offices and Headquarters. A large proportion of the studies identified in this systematic review were conducted in Western European countries, with a relative scarcity of seroprevalence studies from other countries by the end of 2020, an observation noted in other systematic reviews (74-76, 78). This highlights the urgent need for enhanced capacity, the provision of additional support to LMICs and the sharing of information to address the gap in knowledge and tackle research inequity. To counteract the skewedness in the WHO European Region, the WHO Unity protocols have been widely promoted by WHO and ECDC and technical support has been provided to tailor the protocols to local contexts, together with laboratory and financial support to LMICs. In addition, WHO and ECDC jointly established a network of approximately 300 public health professionals to facilitate discussions in related to SARS-CoV-2 seroprevalence, promote timely sharing of results and knowledge and further build capacity in the WHO European Region.

This systematic review is the first in the WHO European region to describe the seroprevalence of SARS-CoV-2 in the first year of the pandemic, prior to the widespread implementation of vaccine programs nationally. In addition, with the inclusion of as yet unpublished data from LMICs, this review contributes to research equity across Member States income levels and provides a more representative overview of the situation in the WHO European Region than published studies alone. This review has some limitations. Firstly, there was significant heterogeneity among the studies, including sampling frame, population and stage of epidemic at time of serosurvey, which makes comparability across studies difficult. Due to such heterogeneity, we opted to not provide one pooled estimate nor conduct a meta-analysis as interpretation would be difficult and may not accurately reflect the picture in the WHO European Region. Secondly, while population-based serological surveys can provide a more accurate estimation of the overall rates of SARS-CoV-2 infection within a population, this approach does not consider antibody waning, which cannot be easily accounted for as antibody levels vary depending on disease severity (85) and longevity is expected to vary greatly across SARS-CoV-2 infected individuals (86). Finally, due to the rapid accumulation of data related to SARS-CoV-2 seroepidemiology and the advent of the ‘preprint era’, not all included studies have been published and may therefore be subject to change upon peer review.

## Conclusion

As SARS-CoV-2 continues to circulate widely, understanding the population seropositivity remains critical for policymakers and public health officials to make informed decisions on optimal public health interventions, such as lifting or tightening of restrictions (9, 87). We found evidence that SARS-CoV-2 antibody seroprevalence across the WHO European Region was low prior to widespread circulation and remained low in the general population during 2020. This suggests that much of the population remained susceptible to infection prior to the implementation of national COVID-19 vaccine campaigns from early 2021 onwards. We also found variation in seroprevalence estimates between and within countries during 2020 with evidence of increased prevalence in areas following high levels of transmission and some association with incidence and mortality trends over time. It is clear that antibody-mediated ‘herd immunity’ through natural infection is not attainable in most countries and COVID-19 vaccines should continue to be distributed widely and equitably to protect priority groups and the wider population. In addition, all efforts must be also directed towards well-informed and evidence-based implementation and maintenance of non-pharmaceutical interventions at a local and national level to stem any future waves of the pandemic. As vaccine programs continue to be implemented more widely, seroprevalence studies will be instrumental to evaluate both natural and vaccine derived immunity overtime to guide public health actions and decision making.

## Data Availability

The unpublished data supporting the findings of this study are available on the open source Zenodo repository https://zenodo.org/communities/unity-sero-2021?page=1&size=20.

## Acknowledgements

The authors would like to acknowledge the Principle Investigators of Unity-aligned studies who have kindly shared their unpublished data: Bosnia and Herzegovina: Dejan Bokonjić (University of East Sarajevo Faculty of Medicine Foča, Bosnia and Herzegovina (Republic of Srpska)), Elma Catovic Baralija (Institute for Transfusion Medicine of the Federation of Bosnia and Herzegovina, Sarajevo, Bosnia and Herzegovina), Sanjin Musa (Institute for Public Health of the Federation of Bosnia and Herzegovina, Sarajevo, Bosnia and Herzegovina (Federation of Bosnia and Herzegovina)), Ranko Škrbić (Medical Faculty in Banja Luka, Bosnia and Herzegovina (Republic of Srpska)); Bulgaria: Angel Kunchev (Ministry of Health of the Republic of Bulgaria), Savina Stoitsova (National Center of Infectious and Parasitic Diseases); Georgia: Olgha Tarkhan-Mouravi (National Center for Disease Control and Public Health Georgia, Head of Vaccine-Preventable and Respiratory Diseases Division, Georgia); Kyrgyzstan: Tatyana Kuchuk (Republican Scientific and Practical Center for Laboratory Diagnostic Quality Control of infectious diseases, Ministry of Health of the Kyrgyz Republic, Kyrgyzstan), Nurmatov Zuridin (Republican Scientific and Practical Center for Control of Viral Infections, Ministry of Health of the Kyrgyz Republic, Kyrgyzstan); Republic of Moldova: Alexei Ceban (National Agency for Public Health, Chişinău, Republic of Moldova). We would also like to thank Céline Roman (WHO EURO), Jeffrey Pires (WHO EURO) and Tommi Karki (ECDC) for data analysis and visualization; Tomas Allen and the WHO HQ library team for development of the search strategy; SeroTracker (Mairead Whelan, Zihan Li, Niklas Bobrovitz and Rahul K Arora) and WHO HQ colleagues (Hannah Lewis and Brianna Cheng); and all ECDC, WHO Headquarters, Regional and Country Office colleagues who contributed to this work. The authors are alone responsible for the views expressed in this publication and they do not necessarily represent the decisions or policies of the WHO or ECDC.

## References

1. World Health Organization; 2021. WHO Coronavirus Disease (COVID-19) Dashboard. Available from: https://covid19.who.int/.

2. Bergeri I, Lewis HC, Subissi L, Nardone A, Valenciano M, Cheng B, et al. Early epidemiological investigations: World Health Organization UNITY protocols provide a standardized and timely international investigation framework during the COVID-19 pandemic. Influenza Other Respir Viruses. 2021;1–7..

3. Cheng MP, Papenburg J, Desjardins M, Kanjilal S, Quach C, Libman M, et al. Diagnostic Testing for Severe Acute Respiratory Syndrome-Related Coronavirus 2: A Narrative Review. Ann Intern Med. 2020;172(11):726–34.

4. Byambasuren O, Cardona M, Bell K, Clark J, McLaws M-L, Glasziou P. Estimating the extent of asymptomatic COVID-19 and its potential for community transmission: Systematic review and meta-analysis. Official Journal of the Association of Medical Microbiology and Infectious Disease Canada. 2020;5(4):223–34.

5. Koh WC, Naing L, Chaw L, Rosledzana MA, Alikhan MF, Jamaludin SA, et al. What do we know about SARS-CoV-2 transmission? A systematic review and meta-analysis of the secondary attack rate and associated risk factors. PLOS ONE. 2020;15(10):e0240205.

6. Buitrago-Garcia D, Egli-Gany D, Counotte MJ, Hossmann S, Imeri H, Ipekci AM, et al. Occurrence and transmission potential of asymptomatic and presymptomatic SARS-CoV-2 infections: A living systematic review and meta-analysis. PLoS Med. 2020;17(9):e1003346.

7. Tanne JH. Covid-19: US cases are greatly underestimated, seroprevalence studies suggest. BMJ. 2020;370:m2988.

8. Wajnberg A, Mansour M, Leven E, Bouvier NM, Patel G, Firpo-Betancourt A, et al. Humoral response and PCR positivity in patients with COVID-19 in the New York City region, USA: an observational study. The Lancet Microbe. 2020;1(7):e283–e9.

9. Murhekar MV, Clapham H. COVID-19 serosurveys for public health decision making. The Lancet Global Health. 2021;9(5):e559–e60.

10. Page MJ, McKenzie JE, Bossuyt PM, Boutron I, Hoffmann TC, Mulrow CD, et al. The PRISMA 2020 statement: an updated guideline for reporting systematic reviews. BMJ. 2021;372:71.

11. Ouzzani M, Hammady H, Fedorowicz Z, Elmagarmid A. Rayyan — a web and mobile app for systematic reviews. 2016;5(210).

12. Van Walle I, Leitmeyer K, Broberg EK. Meta-analysis of the clinical performance of commercial SARS-CoV-2 nucleic acid, antigen and antibody tests up to 22 August 2020. medRxiv. 2020:2020.09.16.20195917.

13. Gudbjartsson DF, Norddahl GL, Melsted P, Gunnarsdottir K, Holm H, Eythorsson E, et al. Humoral Immune Response to SARS-CoV-2 in Iceland. N Engl J Med. 2020;383(18):1724–34.

14. Koronaepidemian väestöserologiatutkimuksen viikkoraportti. Finnish Institute for Health and Welfare – THL 2021. 01/01/2021. Available from: https://www.thl.fi/roko/cov-vaestoserologia/sero_report_weekly.html

15. Zakhashvili K. Sero-epidemiological Cross-sectional investigation on COVID-19 virus infection in Georgia. 2020.

16. Roederer T, Mollo B, Vincent C, Nikolay B, Llosa AE, Nesbitt R, et al. Seroprevalence and risk factors of exposure to COVID-19 in homeless people in Paris, France: a cross-sectional study. The Lancet Public Health. 2021;6(4):e202–e9.

17. Runkel S, Kowalzik F, Gehring S, Winter J, Grandt CL, Marron M, et al. Prevalence of Severe Acute Respiratory Syndrome Coronavirus-2-specific Antibodies in German Blood Donors during the COVID-19 Pandemic. Clin lab. 2020;66(10).

18. Lundkvist Å, Hanson S, Olsen B. Pronounced difference in Covid-19 antibody prevalence indicates cluster transmission in Stockholm, Sweden. Infection Ecology & Epidemiology. 2020;10(1):1806505.

19. Egerup P, Fich Olsen L, Christiansen A-MH, Westergaard D, Severinsen ER, Hviid KVR, et al. Severe Acute Respiratory Syndrome Coronavirus 2 (SARS-CoV-2) Antibodies at Delivery in Women, Partners, and Newborns. Obstetrics & Gynecology. 2021;137(1):49–55.

20. Mattern J, Vauloup-Fellous C, Zakaria H, Benachi A, Carrara J, Letourneau A, et al. Post lockdown COVID-19 seroprevalence and circulation at the time of delivery, France. PLoS One. 2020;15(10):e0240782–e.

21. Tsatsaris V, Mariaggi A-A, Launay O, Couffignal C, Rousseau J, Ancel PY, et al. SARS-COV-2 IgG antibody response in pregnant women at delivery. J Gynecol Obstet Hum Reprod. 2020;50(7):102041-.

22. Cosma S, Borella F, Carosso A, Sciarrone A, Cusato J, Corcione S, et al. The “scar” of a pandemic: Cumulative incidence of COVID-19 during the first trimester of pregnancy. Journal of Medical Virology. 2020;93(1):537–40.

23. Crovetto F, Crispi F, Llurba E, Figueras F, Gómez-Roig MD, Gratacós E. SEROPREVALENCE AND CLINICAL SPECTRUM OF SARS-CoV-2 INFECTION IN THE FIRST VERSUS THIRD TRIMESTER OF PREGNANCY. medRxiv. 2020:2020.06.17.20134098.

24. VillalaÍn C, Herraiz I, Luczkowiak J, Pérez-Rivilla A, Folgueira MD, MejÍa I, et al. Seroprevalence analysis of SARS-CoV-2 in pregnant women along the first pandemic outbreak and perinatal outcome. PLoS One. 2020;15(11):e0243029–e.

25. Lumley SF, Eyre DW, McNaughton AL, Howarth A, Hoosdally S, Hatch SB, et al. SARS-CoV-2 antibody prevalence, titres and neutralising activity in an antenatal cohort, United Kingdom, 14 April to 15 June 2020. Euro surveill. 2020;25(42).

26. Castro Dopico X, Muschiol S, Christian M, Hanke L, Sheward DJ, Grinberg NF, et al. Seropositivity in blood donors and pregnant women during the first year of SARS-CoV-2 transmission in Stockholm, Sweden. Journal of Internal Medicine. 2021;290(3):666–76.

27. Emmenegger M, De Cecco E, Lamparter D, Jacquat RlPB, Ebner D, Schneider MM, et al. Early peak and rapid decline of SARS-CoV-2 seroprevalence in a Swiss metropolitan region. medRxiv. 2020:2020.05.31.20118554-2020.05.31.

28. Clarke C, Prendecki M, Dhutia A, Ali MA, Sajjad H, Shivakumar O, et al. High Prevalence of Asymptomatic COVID-19 Infection in Hemodialysis Patients Detected Using Serologic Screening. J Am Soc Nephrol. 2020;31(9):1969–75.

29. Zambelli A, Fotia V, Bosetti T, Negrini G, di Croce A, Moro C, et al. Prevalence and clinical impact of asymptomatic or mildly symptomatic SARSCoV-2 infection among actively treated cancer patients during COVID-19 pandemic in Italy. Annals of Oncology. 2020;31:S994.

30. Cabezón-Gutiérrez L, Custodio-Cabello S, Palka-Kotlowska M, Oliveros-Acebes E, GarcÍa-Navarro MJ, Khosravi-Shahi P. Seroprevalence of SARS-CoV-2-specific antibodies in cancer outpatients in Madrid (Spain): A single center, prospective, cohort study and a review of available data. Cancer Treat Rev. 2020;90:102102-.

31. Montenegro P, Brotons C, Serrano J, FernÃ¡ndez D, Garcia-Ramos C, Ichazo Ba, et al. Community seroprevalence of COVID-19 in probable and possible cases at primary health care centres in Spain. 2020.

32. Stringhini S, Wisniak A, Piumatti G, Azman AS, Lauer SA, Baysson H, et al. Seroprevalence of anti-SARS-CoV-2 IgG antibodies in Geneva, Switzerland (SEROCoV-POP): a population-based study. The Lancet. 2020;396(10247):313–9.

33. Vos ERA, den Hartog G, Schepp RM, Kaaijk P, van Vliet J, Helm K, et al. Nationwide seroprevalence of SARS-CoV-2 and identification of risk factors in the general population of the Netherlands during the first epidemic wave. J epidemiol community health (1979). 2020.

34. Zuridin; N, Tatyana K. Population-based age-stratified seroepidemiological investigation protocol for coronavirus 2019 (COVID-19) infection in Kyrgyz Republic. 2020.

35. Pedersen OB, Nissen J, Dinh KM, Schwinn M, Kaspersen KA, Boldsen JK, et al. SARS-CoV-2 infection fatality rate among elderly retired Danish blood donors - A cross-sectional study. Clin infect dis. 2020.

36. Stefanelli P, Bella A, Fedele G, Pancheri S, Leone P, Vacca P, et al. Prevalence of SARS-CoV-2 IgG antibodies in an area of northeastern Italy with a high incidence of COVID-19 cases: a population-based study. Clin microbiol infect. 2020.

37. Barchuk A, Skougarevskiy D, Titaev K, Shirokov D, Raskina Y, Novkunkskaya A, et al. Seroprevalence of SARS-CoV-2 antibodies in Saint Petersburg, Russia: a population-based study. Scientific Reports. 2021;11(1):12930.

38. Richard A, Wisniak A, Perez-Saez J, Garrison-Desany H, Petrovic D, Piumatti G, et al. Seroprevalence of anti-SARS-CoV-2 IgG antibodies, risk factors for infection and associated symptoms in Geneva, Switzerland: a population-based study. medRxiv. 2020:2020.12.16.20248180.

39. Jogi P, Soeorg H, Ingerainen D, Soots M, Lattekivi F, Naaber P, et al. Seroprevalence of SARS-CoV-2 IgG antibodies in two regions of Estonia (KoroSero-EST-1). 2020.

40. Sanjin Musa, Elma Catovic Baralija, Mia Blazevic, Seila Cilovic-Lagarija, Gorana Ahmetovic-Karic, Alma Ljuca, et al. Seroepidemiological investigation for coronavirus 2019 (COVID-19) infection in the Federation of Bosnia and Herzegovina. 2020.

41. Tsertsvadze T, Gatserelia L, Mirziashvili M, Dvali N, Abutidze A, Metchurtchlishvili R, et al. SARS-CoV-2 antibody seroprevalence in Tbilisi, the capital city of country of Georgia. medRxiv. 2020:2020.09.18.20195024.

42. Tsitsilonis OE, Paraskevis D, Lianidou E, Pierros V, Akalestos A, Kastritis E, et al. Seroprevalence of Antibodies against SARS-CoV-2 among the Personnel and Students of the National and Kapodistrian University of Athens, Greece: A Preliminary Report. Life-Basel. 2020;10(9).

43. Pagani G, Conti F, Giacomelli A, Bernacchia D, Rondanin R, Prina A, et al. Seroprevalence of SARS-CoV-2 significantly varies with age: Preliminary results from a mass population screening. Journal of Infection. 2020;81(6):E10–E2.

44. Fiore JR, Centra M, De Carlo A, Granato T, Rosa A, Sarno M, et al. Results from a survey in healthy blood donors in South Eastern Italy indicate that we are far away from herd immunity to SARS-CoV-2. J med virol. 2020.

45. Vena A, Berruti M, Adessi A, Blumetti P, Brignole M, Colognato R, et al. Prevalence of antibodies to SARS-CoV-2 in Italian adults and associated risk factors. J Clin Med. 2020;9(9):1–9.

46. Gallian P, Pastorino B, Morel P, Chiaroni J, Ninove L, de Lamballerie X. Lower prevalence of antibodies neutralizing SARS-CoV-2 in group O French blood donors. Antiviral Res. 2020;181:104880-.

47. Ward H, Cooke G, Atchison C, Whitaker M, Elliott J, Moshe M, et al. Declining prevalence of antibody positivity to SARS-CoV-2: a community study of 365,000 adults. medRxiv. 2020:2020.10.26.20219725.

48. Slot E, Hogema BM, Reusken Reimerink JHC, Molier M, Karregat JHM, J NovotnÃ1/2 VMJIJ, et al. Low SARS-CoV-2 seroprevalence in blood donors in the early COVID-19 epidemic in the Netherlands. Nat Commun. 2020;11(1):5744-.

49. Popova Ezhlova, Yu EBA, Melnikova AA, Stepanova TF, Sharukho GV, Letyushev AN, et al. Distribution of SARS-CoV-2 seroprevalence among residents of the Tyumen Region during the COVID-19 epidemic period. Journal of microbiology, epidemiology and immunobiology. Journal of Microbiology Epidemiology Immunobiology. 2020;97(5):392–400.

50. Knabl L, Mitra T, Kimpel J, Rössler A, Volland A, Walser A, et al. High SARS-CoV-2 seroprevalence in children and adults in the Austrian ski resort of Ischgl. Communications Medicine. 2021;1(1):4.

51. Vince A, Zadro R, Sostar Z, Sternak SL, Vranes J, Skaro V, et al. SARS-CoV-2 Seroprevalence in a Cohort of Asymptomatic, RT-PCR Negative Croatian First League Football Players. 2020.

52. Fontanet A, Tondeur L, Grant R, Temmam S, Madec Y, Bigot T, et al. SARS-CoV-2 infection in schools in a northern French city: a retrospective serological cohort study in an area of high transmission, France, January to April 2020. Eurosurveillance. 2021;26(15):2001695.

53. Streeck H, Schulte B, Kummerer BM, Richter E, Holler T, Fuhrmann C, et al. Infection fatality rate of SARS-CoV2 in a super-spreading event in Germany. Nat Commun. 2020;11(1):5829.

54. Santos-Hövener C, Neuhauser HK, Rosario AS, Busch M, Schlaud M, Hoffmann R, et al. Serology- and PCR-based cumulative incidence of SARS-CoV-2 infection in adults in a successfully contained early hotspot (CoMoLo study), Germany, May to June 2020. Euro surveill. 2020;25(47).

55. Pérez-GarcÍa F, Pérez-Zapata A, Arcos N, De la Mata M, Ortiz M, Simón E, et al. Severe acute respiratory coronavirus virus 2 (SARS-CoV-2) infection among hospital workers in a severely affected institution in Madrid, Spain: A surveillance cross-sectional study. Infection Control & Hospital Epidemiology. 2021;42(7):803–9.

56. Herzog S, De Bie J, Abrams S, Wouters I, Ekinci E, Patteet L, et al. Seroprevalence of IgG antibodies against SARS coronavirus 2 in Belgium – a serial prospective cross-sectional nationwide study of residual samples (March – October 2020). medRxiv. 2021:2020.06.08.20125179.

57. Roxhed N, Bendes A, Dale M, Mattsson C, Hanke L, Dodig-Crnković T, et al. Multianalyte serology in home-sampled blood enables an unbiased assessment of the immune response against SARS-CoV-2. Nature Communications. 2021;12(1):3695.

58. Valenti L, Bergna A, Pelusi S, Facciotti F, Lai A, Tarkowski M, et al. SARS-CoV-2 seroprevalence trends in healthy blood donors during the COVID-19 Milan outbreak.

59. Armann JP, Unrath M, Kirsten C, Lueck C, Dalpke A, Berner R. Anti-SARS-CoV-2 IgG antibodies in adolescent students and their teachers in Saxony, Germany (SchoolCoviDD19): very low seropraevalence and transmission rates.

60. Mack D, Gartner BC, Rossler A, Kimpel J, Donde K, Harzer O, et al. Prevalence of SARS-CoV-2 IgG antibodies in a large prospective cohort study of elite football players in Germany (May-June 2020): implications for a testing protocol in asymptomatic individuals and estimation of the rate of undetected cases. Clin Microbiol Infect. 2021;27(3):473 e1–e4.

61. Le Vu S, Jones G, Anna F, Rose T, Richard J-B, Bernard-Stoecklin S, et al. Prevalence of SARS-CoV-2 antibodies in France: results from nationwide serological surveillance. Nature Communications. 2021;12(1):3025.

62. COVID-19: The National Prevalence Survey. Results of antibody test with 18,000 invited participants, week 34-36.. Statens Serum Institute; 2020 07/10/2020. 07/10/2020. Accessed: Available from: https://www.ssi.dk/-/media/arkiv/dk/aktuelt/nyheder/2020/notat---covid-19-prvalensundersgelsen.pdf?la=da

63. Lenicek K J.,, Zrinski Topic R, Stevanovic V, Lukic-Grlic A, Tabain I, Misak Z, et al. Seroprevalence of SARS-CoV-2 infection among children in Children’s Hospital Zagreb during the initial and second wave of COVID-19 pandemic in Croatia. (1846-7482 (Electronic)).

64. Enhanced Surveillance of COVID-19 in Scotland: Population-based seroprevalence surveillance Public Health Scotland; 2020 30 September 2020. 30 September 2020. Accessed:01/01/2021. Available from: https://publichealthscotland.scot/publications/enhanced-surveillance-of-covid-19-in-scotland/enhanced-surveillance-of-covid-19-in-scotland-population-based-seroprevalence-surveillance-30-september-2020/

65. Sweden PHAo. PÅvisning av antikroppar efter genomgÅngen COVID-19 i blodprov frÅn öppenvÅrden (Delrapport 1). 2020. Available from: https://www.folkhalsomyndigheten.se/publicerat-material/publikationsarkiv/p/pavisning-av-antikroppar-efter-genomgangen-covid-19-i-blodprov-fran-oppenvarden-delrapport-1/

66. PÅvisning av antikroppar efter genomgÅngen COVID-19 hos blodgivare (Delrapport 2). Public Health Agency of Sweden (Folkhälsomyndigheten); 2020. 01/06/2021. Available from: https://www.folkhalsomyndigheten.se/publicerat-material/publikationsarkiv/p/pavisning-av-antikroppar-efter-genomgangen-covid-19-hos-blodgivare-delrapport-2/

67. Weekly Coronavirus Disease 2019 (COVID-19) Surveillance Report: Week 22. Public Health England; 2020. 01/01/2021. Available from: https://assets.publishing.service.gov.uk/government/uploads/system/uploads/attachment_data/file/888254/COVID19_Epidemiological_Summary_w22_Final.pdf

68. Weekly Coronavirus Disease 2019 (COVID-19) Surveillance Report: Week 39. Public Health England; 2020. 01/01/2021. Available from: https://assets.publishing.service.gov.uk/government/uploads/system/uploads/attachment_data/file/921561/Weekly_COVID19_Surveillance_Report_week_39_FINAL.pdf

69. Weekly Coronavirus Disease 2019 (COVID-19) surveillance report: Week 38. Public Health England; 2020. Available from: https://assets.publishing.service.gov.uk/government/uploads/system/uploads/attachment_data/file/919676/Weekly_COVID19_Surveillance_Report_week_38_FINAL_UPDATED.pdf

70. Weekly Coronavirus Disease 2019 (COVID-19) surveillance report: Week 37. Public Health England; 2020. 01/01/2021. Available from: https://assets.publishing.service.gov.uk/government/uploads/system/uploads/attachment_data/file/920372/Weekly_COVID19_Surveillance_Report_week_37_FINAL_UPDATED.pdf

71. Weekly Coronavirus Disease 2019 (COVID-19) surveillance report: Week 36. Public Health England; 2020. 01/01/2021. Available from: https://assets.publishing.service.gov.uk/government/uploads/system/uploads/attachment_data/file/920373/Weekly_COVID19_Surveillance_Report_week_36_UPDATED.pdf

72. Weekly Coronavirus Disease 2019 (COVID-19) surveillance report: Week 35. Public Health England; 2020. 01/01/2021. Available from: https://assets.publishing.service.gov.uk/government/uploads/system/uploads/attachment_data/file/912973/Weekly_COVID19_Surveillance_Report_week_35_FINAL.PDF

73. Ulyte A, Radtke T, Abela IA, Haile SR, Berger C, Huber M, et al. Clustering and longitudinal change in SARS-CoV-2 seroprevalence in school children in the canton of Zurich, Switzerland: prospective cohort study of 55 schools. BMJ. 2021;372:n616.

74. Bobrovitz N, Arora RK, Cao C, Boucher E, Liu M, Donnici C, et al. Global seroprevalence of SARS-CoV-2 antibodies: A systematic review and meta-analysis. PLOS ONE. 2021;16(6):e0252617.

75. Rostami A, Sepidarkish M, Leeflang MMG, Riahi SM, Nourollahpour Shiadeh M, Esfandyari S, et al. SARS-CoV-2 seroprevalence worldwide: a systematic review and meta-analysis. Clinical Microbiology and Infection. 2021;27(3):331–40.

76. Chen X, Chen Z, Azman AS, Deng X, Sun R, Zhao Z, et al. Serological evidence of human infection with SARS-CoV-2: a systematic review and meta-analysis. The Lancet Global Health. 2021;9(5):e598–e609.

77. Bobrovitz N, Arora RK, Cao C, Boucher E, Liu M, Rahim H, et al. Global seroprevalence of SARS-CoV-2 antibodies: a systematic review and meta-analysis. medRxiv. 2020:2020.11.17.20233460.

78. Vaselli NM, Hungerford D, Shenton B, Khashkhusha A, Cunliffe NA, French N. The Seroprevalence of SARS-CoV-2 in Europe: A Systematic Review. bioRxiv. 2021:2021.04.12.439425.

79. Goldman M, Steele WR, Di Angelantonio E, van den Hurk K, Vassallo RR, Germain M, et al. Comparison of donor and general population demographics over time: a BEST Collaborative group study. (1537-2995 (Electronic)).

80. Deeks JJ, Dinnes J, Takwoingi Y, Davenport C, Spijker R, Taylor-Phillips S, et al. Antibody tests for identification of current and past infection with SARS-CoV-2. Cochrane Database of Systematic Reviews. 2020(6).

81. Lisboa Bastos M, Tavaziva G, Abidi SK, Campbell JR, Haraoui L-P, Johnston JC, et al. Diagnostic accuracy of serological tests for covid-19: systematic review and meta-analysis. BMJ. 2020;370:m2516.

82. World Health Organization; 2020. Establishment of the WHO International Standard and Reference Panel for anti-SARS-CoV-2 antibody. Available from: https://www.who.int/publications/m/item/WHO-BS-2020.2403.

83. Kristiansen PA, Page M, Bernasconi V, Mattiuzzo G, Dull P, Makar K, et al. WHO International Standard for anti-SARS-CoV-2 immunoglobulin. The Lancet. 2021;397(10282):1347–8.

84. The JRC releases new reference materials for the quality control of SARS-CoV-2 antibody tests. European Centre for Disease Prevention and Control; 2021. Available from: https://ec.europa.eu/jrc/en/news/new-reference-materials-quality-control-covid-19-antibody-tests

85. Seow J, Graham C, Merrick B, Acors S, Pickering S, Steel KJA, et al. Longitudinal observation and decline of neutralizing antibody responses in the three months following SARS-CoV-2 infection in humans. Nature Microbiology. 2020;5(12):1598–607.

86. Chia WN, Zhu F, Ong SWX, Young BE, Fong S-W, Le Bert N, et al. Dynamics of SARS-CoV-2 neutralising antibody responses and duration of immunity: a longitudinal study. The Lancet Microbe. 2021;2(6):e240–e9.

87. Clapham H Fau - Hay J, Hay J Fau - Routledge I, Routledge I Fau - Takahashi S, Takahashi S Fau - Choisy M, Choisy M Fau - Cummings D, Cummings D Fau - Grenfell B, et al. Seroepidemiologic Study Designs for Determining SARS-COV-2 Transmission and Immunity. (1080-6059 (Electronic)).

88. Wagner A, Guzek A, Ruff J, Jasinska J, Scheikl U, Zwazl I, et al. A longitudinal seroprevalence study in a large cohort of working adults reveals that neutralising SARS-CoV-2 RBD-specific antibodies persist for at least six months independent of the severity of symptoms. medRxiv. 2020:2020.12.22.20248604.

89. Ladage D, Höglinger Y, Ladage D, Adler C, Yalcin I, Braun RJ. SARS-CoV-2 antibody prevalence and symptoms in a local Austrian population. medRxiv. 2020:2020.11.03.20219121.

90. Boey L, Roelants M, Merckx J, Hens N, Desombere I, Duysburgh E, et al. Age-dependent seroprevalence of SARS-CoV-2 antibodies in school-aged children from areas with low and high community transmission. European Journal of Pediatrics. 2021.

91. Bokonjic. A national study of seroprevalence of COVID-19 infection in the population of the Republika Srpska 2020.

92. Kunchev A, Stoitsova S. Seroepidemiologic study to detect antibody-mediated immunity against SARS-CoV-2 among residents and healthcare workers in the city of Plovdiv, Bulgaria, weeks 21-24, 2020. Unpublished2020.

93. De første foreløbige resultater af undersøgelsen for COVID-19 i befolkningen er nu klar. Statens Serum Institut; 2020. 01/01/2021. Available from: https://www.ssi.dk/aktuelt/nyheder/2020/de-forste-forelobige-resultater-af-undersogelsen-for-covid-19-i-befolkningen-er-nu-klar

94. New preliminary results from the representative seroprevalence study of COVID-19. Statens Serum Institute; 2020. 01/01/2021. Available from: https://files.ssi.dk/notat_foreloebige_resultater_pilotundersoegelse_seropraevalens_COVID-19_29_6_2020

95. Petersen MS, Strom M, Christiansen DH, Fjallsbak JP, Eliasen EH, Johansen M, et al. Seroprevalence of SARS-CoV-2-Specific Antibodies, Faroe Islands. Emerg Infect Dis. 2020;26(11):2761–3.

96. Jõgi P, Soeorg H, Ingerainen D, Soots M, Lättekivi F, Naaber P, et al. Seroprevalence of SARS-CoV-2 IgG antibodies in two regions of Estonia (KoroSero-EST-1). medRxiv. 2020:2020.10.21.20216820.

97. Carrat F, de Lamballerie X, Rahib D, Blanché H, Lapidus N, Artaud F, et al. Seroprevalence of SARS-CoV-2 among adults in three regions of France following the lockdown and associated risk factors: a multicohort study. medRxiv. 2020:2020.09.16.20195693.

98. Aziz NA, Corman VM, Echterhoff AKC, Müller MA, Richter A, Schmandke A, et al. Seroprevalence and correlates of SARS-CoV-2 neutralizing antibodies from a population-based study in Bonn, Germany. Nature Communications. 2021;12(1):2117.

99. Weis S, Scherag A, Baier M, Kiehntopf M, Kamradt T, Kolanos S, et al. Antibody response using six different serological assays in a completely PCR-tested community after a coronavirus disease 2019 outbreak—the CoNAN study. Clinical Microbiology and Infection. 2021;27(3):470.e1-.e9.

100. Merkely B, Szabo AJ, Kosztin A, Berenyi E, Sebestyen A, Lengyel C, et al. Novel coronavirus epidemic in the Hungarian population, a cross-sectional nationwide survey to support the exit policy in Hungary. Geroscience. 2020;42(4):1063–74.

101. Guerriero M, Bisoffi Z, Poli A, Micheletto C, Pomari C. Prevalence of asymptomatic SARS-CoV-2-positive individuals in the general population of northern Italy and evaluation of a diagnostic serological ELISA test: a cross-sectional study protocol. BMJ Open. 2020;10(10):e040036–e.

102. Cito F, Amato L, Di Giuseppe A, Danzetta ML, Iannetti S, Petrini A, et al. A COVID-19 Hotspot Area: Activities and Epidemiological Findings. Microorganisms. 2020;8(11):1711-.

103. Snoeck CJ, Vaillant M, Abdelrahman T, Satagopam VP, Turner JD, Beaumont K, et al. Prevalence of SARS-CoV-2 infection in the Luxembourgish population: the CON-VINCE study.

104. Popova Ezhlova, Melânikova AA, Balakhonov SV, Chesnokova MV, Dubrovina VI, Lyalina LV, et al. Experience in studying seroprevalence to SARS-CoV-2 virus in the population of the Irkutsk Region during COVID-19 outbreak. Problemy Osobo Opasnykh Infektsii. 2020(3):106–13.

105. Popova Ezhlova, Melânikova AA, Historik OA, Mosevich OS, Lyalina LV, Smirnov VS, et al. Assessment of the herd immunity to SARS-CoV-2 among the population of the leningrad region during the COVID-19 epidemic. Problemy Osobo Opasnykh Infektsii. 2020(3):114–23.

106. Popova Ezhlova, Melânikova AA, Bashketova NS, Fridman RK, Lyalina LV, Smirnov VS, et al. Herd immunity to SARS-CoV-2 among the population in Saint-Petersburg during the COVID-19 epidemic. Problemy Osobo Opasnykh Infektsii. 2020(3):124–30.

107. Pollan M, Perez-Gomez B, Pastor-Barriuso R, Oteo J, Hernan MA, Perez-Olmeda M, et al. Prevalence of SARS-CoV-2 in Spain (ENE-COVID): a nationwide, population-based seroepidemiological study. Lancet. 2020;396(10250):535–44.

108. Förekomsten av antikroppar mot SARS-CoV-2 i stadsdelen Rinkeby-Kista, Stockholm, 22–24 juni 2020. Public Health Agency of Sweden; 2020 04/09/2020. Report No.: 20129 04/09/2020. Accessed:01/01/2021. Available from: https://www.folkhalsomyndigheten.se/contentassets/2cf102cd299c4382b9a0447dc0626356/forekomsten-antikroppar-rinkeby-kista.pdf

109. Bi Q, Lessler J, Eckerle I, Lauer SA, Kaiser L, Vuilleumier N, et al. Household Transmission of SARS-COV-2: Insights from a Population-based Serological Survey. 2020.

110. Wells PM, Doores KJ, Couvreur S, Nunez RM, Seow J, Graham C, et al. Estimates of the rate of infection and asymptomatic COVID-19 disease in a population sample from SE England. Journal of Infection. 2020.

111. Tsaneva-Damyanova D. SARS-CoV-2: seroepidemiological pattern in northeastern Bulgaria. Biotechnology & Biotechnological Equipment. 2020;34(1):441–6.

112. Bloomfield M, Pospisilova I, Cabelova T, Sediva A, Ibrahimova M, Borecka K, et al. Searching for COVID-19 Antibodies in Czech Childrenâ€”A Needle in the Haystack. Frontiers in Pediatrics. 2020;8.

113. Capai L, Ayhan N, Masse S, Canarelli J, Priet S, Simeoni M-H, et al. Seroprevalence of SARS-CoV-2 IgG Antibodies in Corsica (France), April and June 2020. Journal of Clinical Medicine. 2020;9(11):3569-.

114. Cohen R, Jung C, Ouldali N, Sellam A, Batard C, Cahn-Sellem F, et al. Assessment of spread of SARS-CoV-2 by RT-PCR and concomitant serology in children in a region heavily affected by COVID-19 pandemic. medRxiv. 2020:2020.06.12.20129221.

115. Bogogiannidou Z, Vontas A, Dadouli K, Kyritsi MA, Soteriades S, Nikoulis DJ, et al. Repeated leftover serosurvey of SARS-CoV-2 IgG antibodies, Greece, March and April 2020. Euro surveill. 2020;25(31).

116. Posfay-Barbe KM, Andrey DO, Virzi J, Cohen P, Pigny F, Goncalves AR, et al. Prevalence of IgG against SARS-CoV-2 and evaluation of a rapid MEDsan IgG test in children seeking medical care. Clin infect dis. 2020.

117. Dickson E, Palmateer NE, Murray J, Robertson C, Waugh C, Wallace LA, et al. Enhanced surveillance of COVID-19 in Scotland: population-based seroprevalence surveillance for SARS-CoV-2 during the first wave of the epidemic. Public health. 2021;190:132–4.

118. Erikstrup C, Hother CE, Pedersen OBV, Mølbak K, Skov RL, Holm DK, et al. Estimation of SARS-CoV-2 Infection Fatality Rate by Real-time Antibody Screening of Blood Donors. Clinical Infectious Diseases. 2020;72(2):249–53.

119. Danmark Bi. Bloddonorer testes for overstÅet infektion med coronavirus. Bloddonorerne i Danmark; 2020. 01/01/2021. Available from: www.bloddonor.dk/coronavirus/

120. Grzelak L, Temmam S, Planchais C, Demeret C, Tondeur L, Huon Cl, et al. A comparison of four serological assays for detecting anti-SARS-CoV-2 antibodies in human serum samples from different populations. Sci transl med. 2020;12(559).

121. Fischer B, Knabbe C, Vollmer T. SARS-CoV-2 IgG seroprevalence in blood donors located in three different federal states, Germany, March to June 2020. Euro surveill. 2020;25(28).

122. Percivalle E, Cambiè G, Cassaniti I, Nepita EV, Maserati R, Ferrari A, et al. Prevalence of SARS-CoV-2 specific neutralising antibodies in blood donors from the Lodi Red Zone in Lombardy, Italy, as at 06 April 2020. Euro surveill. 2020;25(24).

123. Thompson CP, Grayson NE, Paton RS, Bolton JS, LourenÃ§o J, Penman BS, et al. Detection of neutralising antibodies to SARS-CoV-2 to determine population exposure in Scottish blood donors between March and May 2020. Euro surveill. 2020;25(42).

124. Fogel O, Mariaggi AA, Méritet JF, André E, Boisson M, Combier A, et al. Étude prospective de séroprévalence du Sars-Cov-2 chez 249 patients suivis pour un rhumatisme inflammatoire chronique. Revue du Rhumatisme. 2020;87:A292–A3.

125. Choi M, Bachmann F, Naik MG, Duettmann W, Duerr M, Zukunft B, et al. Low Seroprevalence of SARS-CoV-2 Antibodies during Systematic Antibody Screening and Serum Responses in Patients after COVID-19 in a German Transplant Center. J Clin Med. 2020;9(11).

126. Rauber Conrad, Tiwari-Heckler Shilpa, Jan P, Arianeb M, Frederike L, Philip G, et al. SARS-CoV-2 seroprevalence and clinical features of COVID-19 in a German liver transplant recipient cohort: a prospective serosurvey study. Transplantation Proceedings. 2020.

127. Medas F, Cappellacci F, Anedda G, Canu GL, Del Giacco S, Calò PG, et al. Seroprevalence of SARS-Cov-2 in the setting of a non-dedicated COVID-19 hospital in a low CoV-2 incidence area: Implications for surgery. Annals of Medicine and Surgery. 2020.

128. Capasso N, Palladino R, Montella E, Pennino F, Lanzillo R, Carotenuto A, et al. Prevalence of SARS-CoV-2 Antibodies in Multiple Sclerosis: The Hidden Part of the Iceberg. 2020;9(12).

129. Cento V, Alteri C, Merli M, Di Ruscio F, Tartaglione L, Rossotti R, et al. Effectiveness of infection-containment measures on SARS-CoV-2 seroprevalence and circulation from May to July 2020, in Milan, Italy. PLoS One. 2020;15(11):e0242765–e.

130. Berte R, Mazza S, Stefanucci MR, Noviello D, Costa S, Ciafardini C, et al. Seroprevalence of SARS-CoV2 in IBD Patients Treated with Biologic Therapy. J Crohns Colitis. 2021;15(5):864–8.

131. Prados N, GonzÃ¡lez-Ravina C, Vergara V, Herrero J, Cruz M, Requena A. RISK FACTOR ANALYSIS FOR SARS-COV-2 SEROPOSITIVITY WITHIN ASSISTED REPRODUCTIVE. Fertility and Sterility. 2020;114(3):e542–e.

132. Prendecki M, Clarke C, Gleeson S, Greathead L, Santos E, McLean A, et al. Detection of SARS-CoV-2 Antibodies in Kidney Transplant Recipients. J Am Soc Nephrol. 2020;31(12):2753–6.

133. Krátká Z Fau - Fürst T, Fürst T Fau - Vencálek O, Vencálek O Fau - Kůrková V, Kůrková V Fau - Šimečková E, Šimečková E Fau - Fleischmannová J, Fleischmannová J Fau - Strojil J, et al. Exploratory drilling: how to set up, carry out, and evaluate a seroprevalence study. (0008-7335 (Print)).

134. Jerković I, Ljubić T, BaŠić Ž, Kružić I, Kunac N, Bezić J, et al. SARS-CoV-2 Antibody Seroprevalence in Industry Workers in Split-Dalmatia and Šibenik-Knin County, Croatia. J Occup Environ Med. 2021;63(1):32–7.

135. Anna F, Goyard S, Lalanne AI, Nevo F, Gransagne M, Souque P, et al. High seroprevalence but short-lived immune response to SARS-CoV-2 infection in Paris. European Journal of Immunology. 2021;51(1):180–90.

136. Krähling V, Kern M, Halwe S, Müller H, Rohde C, Savini M, et al. Epidemiological study to detect active SARS-CoV-2 infections and seropositive persons in a selected cohort of employees in the Frankfurt am Main metropolitan area. medRxiv. 2020:2020.05.20.20107730-2020.05.20.

137. National Agency for Public Health RoM. Serological surveillance of COVID-19 in the general population with age stratification in the Republic of Moldova. 2020.

138. Gujski M, Jankowski M, Pinkas J, Wierzba W, Samel-Kowalik P, Zaczynski A, et al. Prevalence of Current and Past SARS-CoV-2 Infections among Police Employees in Poland, June-July 2020. Journal of Clinical Medicine. 2020;9(10):11-.

139. Roarty C, Tonry C, McFetridge L, Mitchell H, Watson C, Waterfield T. Kinetics and seroprevalence of SARS-CoV-2 antibodies in children. Lancet, Infect dis (Online). 2020.

140. Waterfield T, Watson C, Moore R, Ferris K, Tonry C, Watt A, et al. Seroprevalence of SARS-CoV-2 antibodies in children: a prospective multicentre cohort study. Arch Dis Child. 2021;106(7):680–6.

141. Reisinger EC, von Possel R, Warnke P, Geerdes-Fenge HF, Hemmer CJ, Pfefferle S, et al. Mütter-Screening in einem COVID-19-Niedrig-Pandemiegebiet: Bestimmung SARS-CoV-2-spezifischer Antikörper bei 401 Rostocker Müttern mittels ELISA und Immunfluoreszenz-Bestätigungstest. Dtsch Med Wochenschr. 2020;145(17):e96–e100.

142. Lastrucci Lorini, Chiara Del Riccio, Marco Gori, Eleonora Chiesi, Fabrizio Sartor, Gino Zanella, et al. SARS-CoV-2 Seroprevalence Survey in People Involved in Different Essential Activities during the General Lock-Down Phase in the Province of Prato (Tuscany, Italy). Vaccines. 2020;8(4):778-.

143. Soriano V, Meiriño R, Corral O, Guallar MP. Severe Acute Respiratory Syndrome Coronavirus 2 Antibodies in Adults in Madrid, Spain. Clin Infect Dis. 2021;72(6):1101–2.

144. Davis KAS, Carr E, Leightley D, Vitiello V, Bergin-Cartwright G, Lavelle G, et al. Indicators of past COVID-19 infection status: Findings from a large occupational cohort of staff and postgraduate research students from a UK university. medRxiv. 2021:2020.12.07.20245183.

