## Supplementary Material for "Seroprevalence of SARS-CoV-2 antibodies prior to the widespread introduction of vaccine programmes in the WHO European Region, January - December 2020: a systematic review"

### S1 Supplementary Methods

#### S1.1 Search strategy and selection criteria

We searched the WHO "COVID-19 Global literature on coronavirus disease" database (MEDLINE, ELSEVIER and the pre-print servers medRxiv and bioRxiv) (100) using search terms that included a range of criteria relating to seroprevalence surveys. The search terms and inclusion and exclusion criteria are described below (Table S1).

#### S1.2 Search terms

ti:sero$surv* or ti:serosurv* or ti:seroepidemiolog* or ti:sero$epidemiolog* or ti:serolog* or ti:seropositiv* or ti:seropositiv* or ti:serosurveillance or ti:sero$surveillance or ti:seroprevalence or ti:sero$prevalence or ti:antibody or ti:antibodies or ti:immunity or ti:immunoglobulin OR ab:sero$surv* or ab:serosurv* or ab:seroepidemiolog* or ab:sero$epidemiolog* or ab:serolog* or ab:seropositiv* or ab:sero$positiv* or ab:serosurveillance or ab:sero$surveillance or ab:seroprevalence or ab:sero$prevalence

### Table S1 Inclusion and exclusion criteria

| Characteristics | Inclusion criteria | Exclusion criteria |
|---|---|---|
| **Type of evidence** | Published or unpublished scientific literature | Media reports (e.g. news items and press releases), reviews, assessed performance of a test, protocols |
| | Completed or ongoing serosurveys | Unrelated to seroprevalence |
| | Report seroprevalence estimates from one or multiple time points | |
| | Cross-sectional and cohort study designs | Case-control studies, case reports or reviews |
| **Population** | Studies of human participants, any age | Studies of non-human participants (i.e animal studies) |
| | Population groups considered to be representative of the general population | Population groups considered to be unrepresentative of the general population as they had higher risk of infection (e.g healthcare workers and other high-risk groups). |
| | | Studies only of individuals with suspected (e.g. respiratory symptoms) or confirmed SARS-CoV-2 (RT-PCR laboratory confirmation) |
| **Geographical location** | WHO European region | Outside of WHO European Region |
| **Languages** | Any language | N/A |

**Table S2: Description of the quality assessment criteria used**

| Risk of bias criteria | Risk of bias assessment | Risk of bias scoring |
|---|---|---|
| **Sampling frame: Representative of general population?** | Poor (e.g hospital admissions, GP visits, pregnant women, employees) | 0 |
| | Weak (eg. blood donors) | 1 |
| | Good (e.g household sampling) | 2 |
| **Sampling frame: Age profile included?** | Does not cover those in target population (e.g study of 'all ages' excludes elderly or children) | 0 |
| | Study includes all those in target population | 1 |
| | Includes all ages (including children) | 2 |
| **Sampling method: Were study participants sampled appropriate?** | Non-random/non-exhaustive (including convenience sampling) | 0 |
| | Exhaustive | 1 |
| | Random | 2 |
| **Sample size: Is sample size calculation described in methods?** | Unclear | 0 |
| | Yes | 1 |
| **Sample size: Was the sample size adequate?** | If at least 300 samples in the study then adequate OR if at least 100 samples per age group (if stratified by age) | 1 |
| | If NONE of above OR no mention in methods/unclear | 0 |
| **Test method: Use of more than one assays/test?** | Yes | 1 |
| | No | 0 |
| **Test method 1: Are tests sufficiently accurate? (no clinical validation)** | Sensitivity/specificity acceptable* (i.e ELISA: Sn ≥95%, Sp >97% OR POCT: Sn ≥90%, Sp >97%) | 1 |
| | Sensitivity/specificity not acceptable (i.e ELISA: Sn <95%, Sp <97% OR POCT: Sn <90%, Sp <97%) | 0 |
| **Test method 2: Use of commercial tests with clinical validation?** | Sensitivity/specificity acceptable* (i.e ELISA: Sn ≥95%, Sp >97% OR POCT: Sn ≥90%, Sp >97%) | 2 |
| | Sensitivity/specificity not acceptable (i.e ELISA: Sn <95%, Sp <97% OR POCT: Sn <90%, Sp <97%) | 1 |
| **Test method 3: Use of in-house assay** | Sensitivity/specificity acceptable* (i.e ELISA: Sn ≥95%, Sp >97% OR POCT: Sn ≥90%, Sp >97%) | 2 |
| | Sensitivity/specificity not acceptable (i.e ELISA: Sn <95%, Sp <97% OR POCT: Sn <90%, Sp <97%) | 1 |
| | No results reported | 0 |
| **Data analysis: Were the results adjusted according to sensitivity/specificity of the serological test?** | Yes and Confidence intervals presented | 2 |
| | Yes but no Confidence intervals presented | 1 |
| | No/Unclear | 0 |
| **Overall risk of bias** | **High risk of bias** | **1-3** |
| | **Moderate risk of bias** | **4-6** |
| | **Low-risk bias** | **>6** |

### S2 Supplementary Results

**Figure S1 Forest plot of the seroprevalence of SARS-CoV-2 antibodies in Community/Household samples with corresponding 95% confidence intervals in WHO European Region (1/1/2020-31/12/2020)**

See PDF file attached separately

**Figure S2: Forest plot of the seroprevalence of SARS-CoV-2 antibodies in (Top to bottom, Left to Right) a) Residual sera b) Blood donors c) Pregnant or Parturient women d) Patients seeking care (non-COVID) and e) Other populations**

See PDF file attached separately

**Figure S3 Time point of conducted sero-epidemiology studies in relation to country epidemic activity**

See PDF file attached separately
