## Supplementary figures and images for "Seroprevalence of SARS-CoV-2 antibodies prior to the widespread introduction of vaccine programmes in the WHO European Region, January - December 2020: a systematic review"

### Supplementary S1

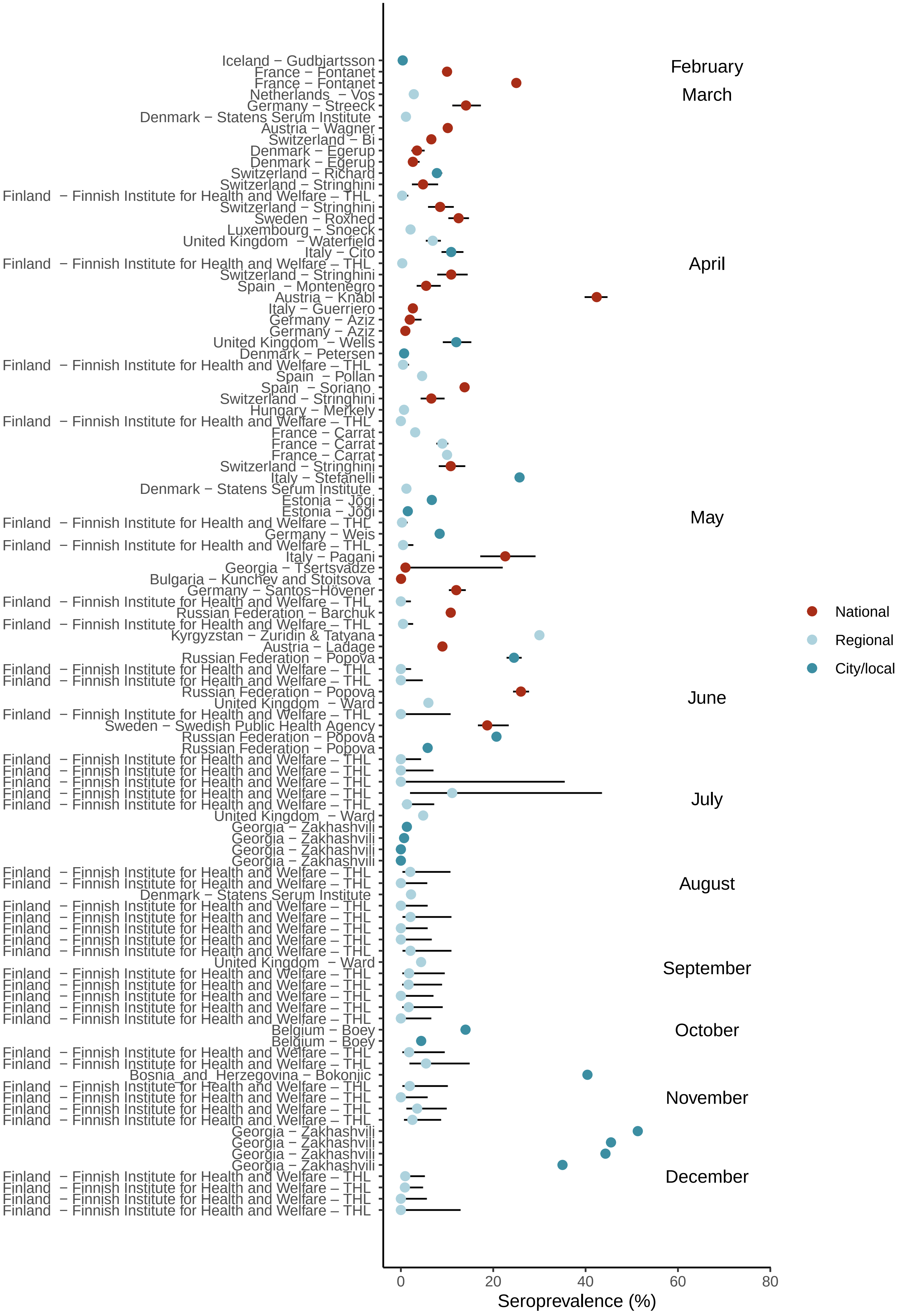

### Supplementary S2

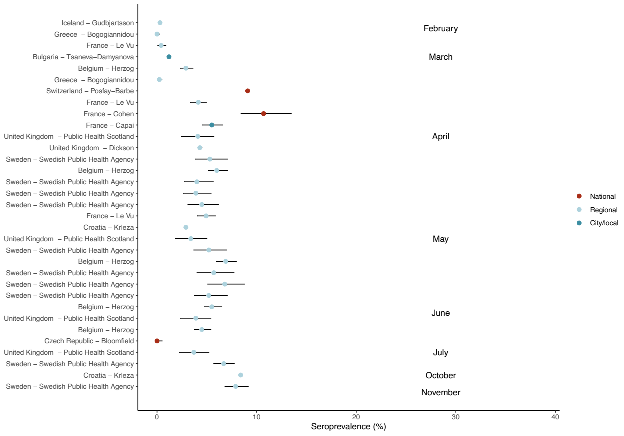

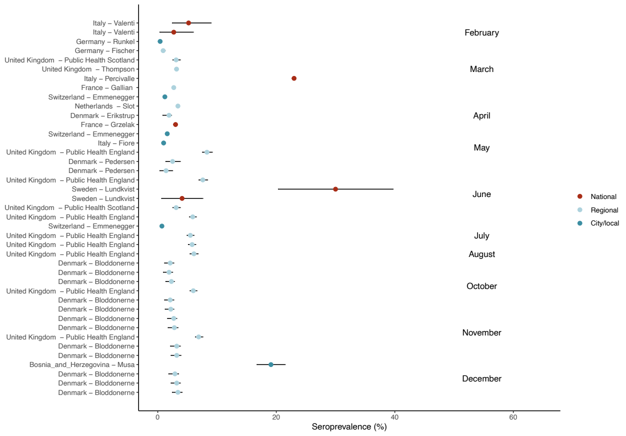

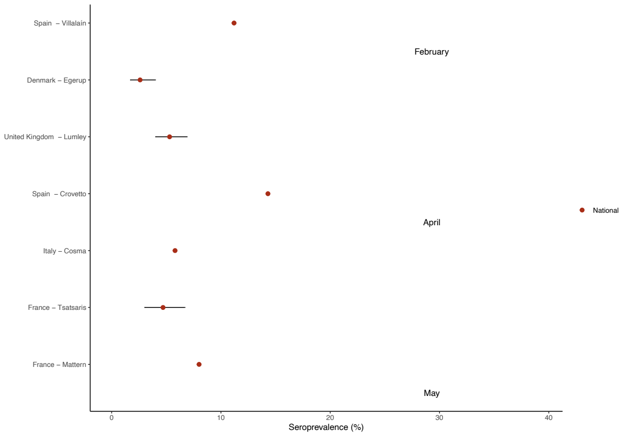

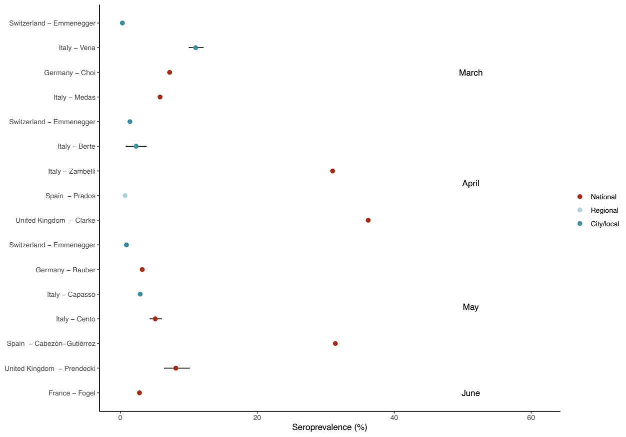

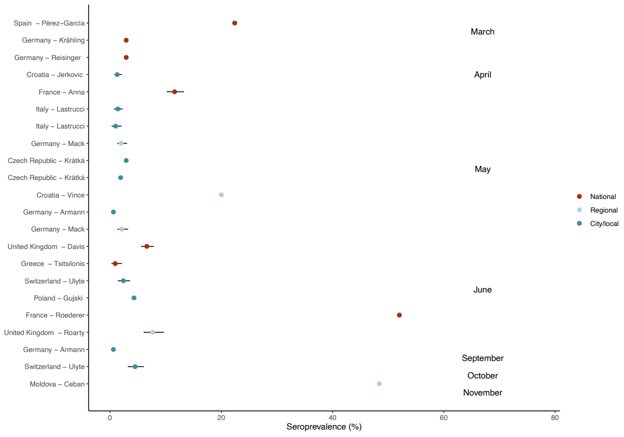

### Supplementary S3

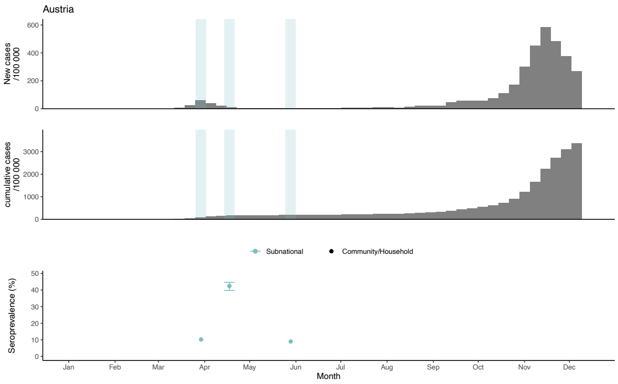

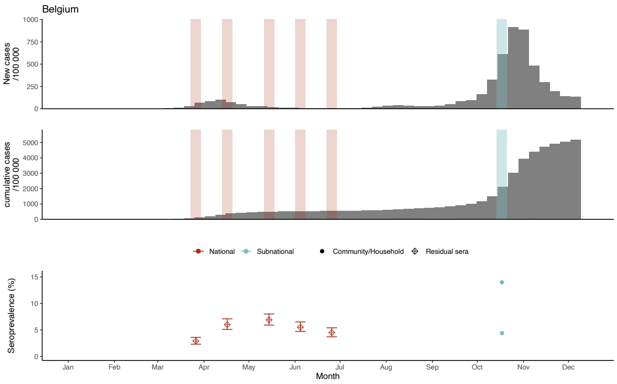

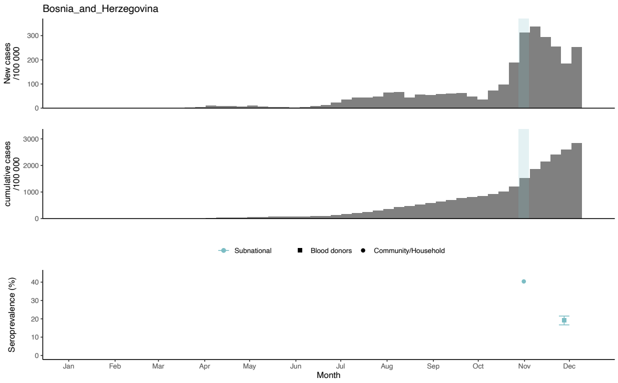

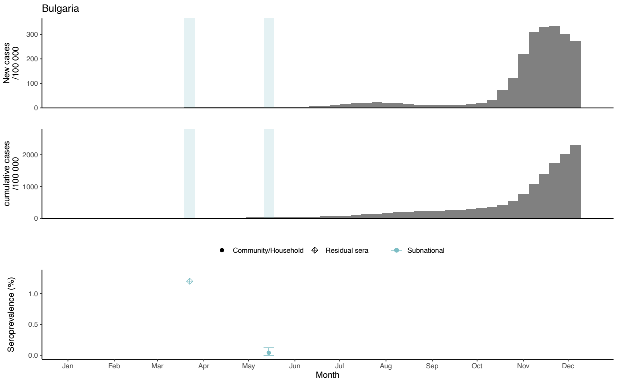

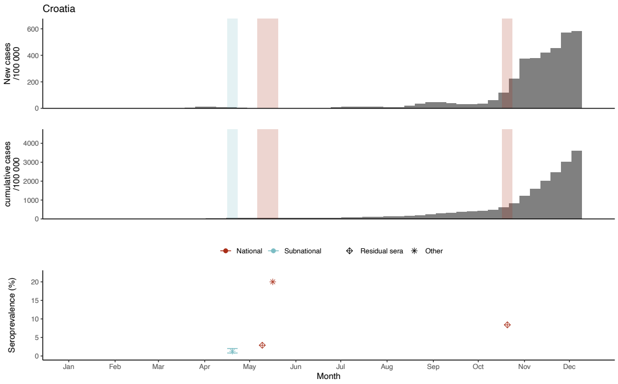

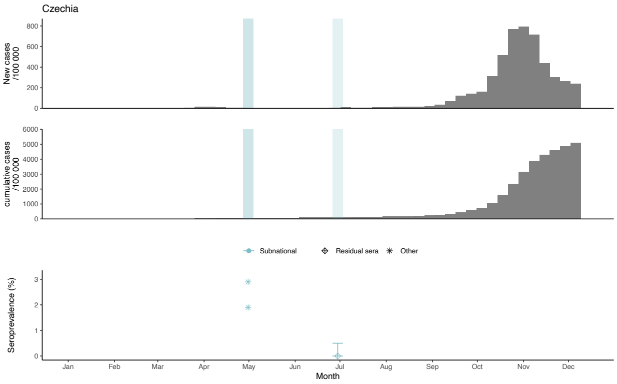

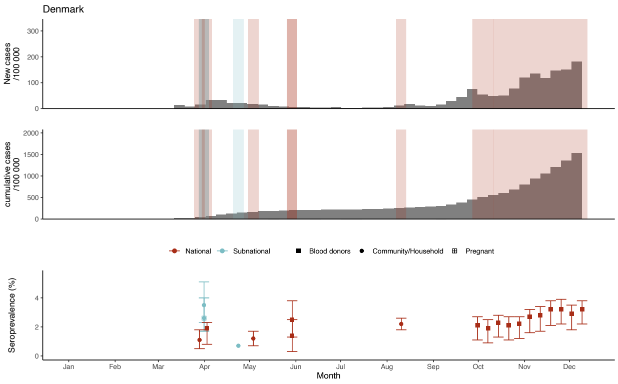

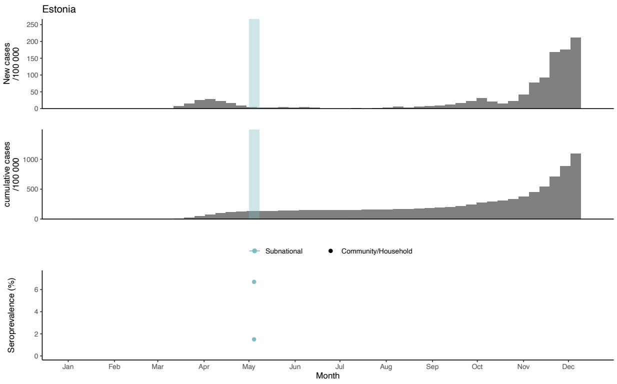

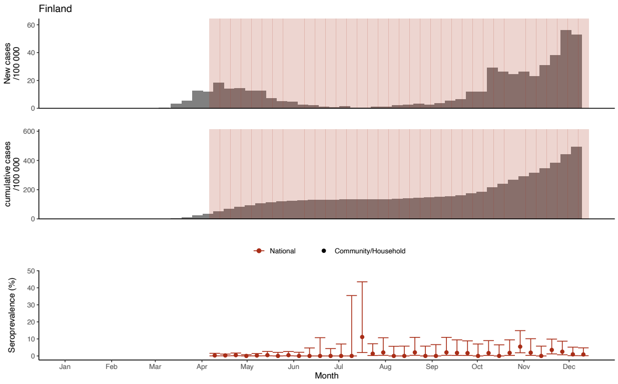

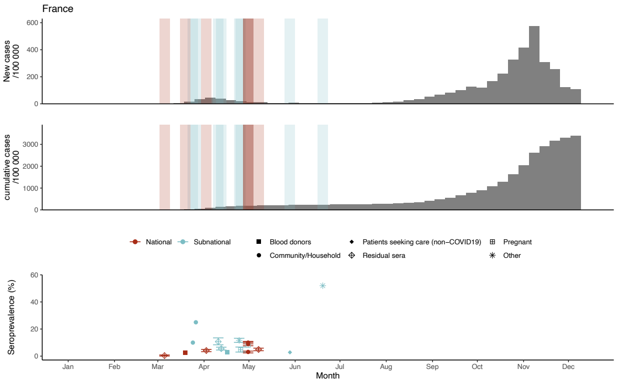

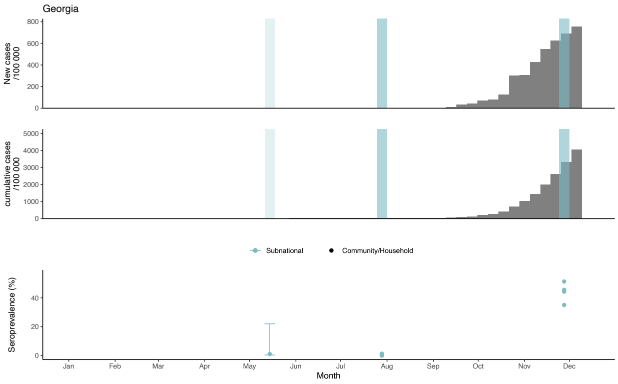

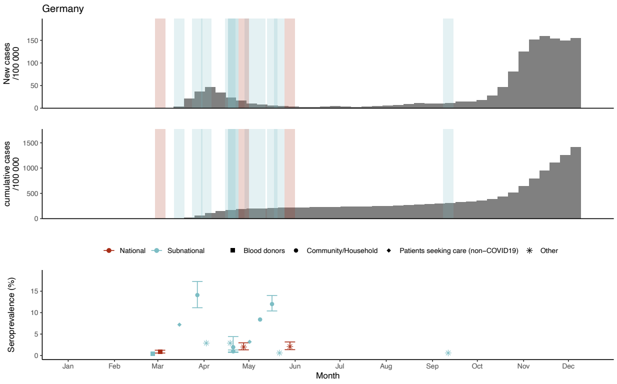

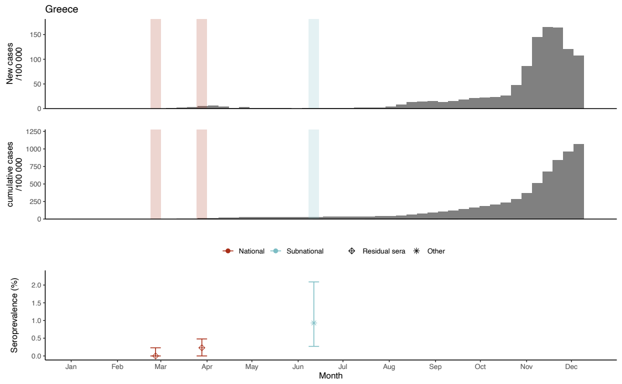

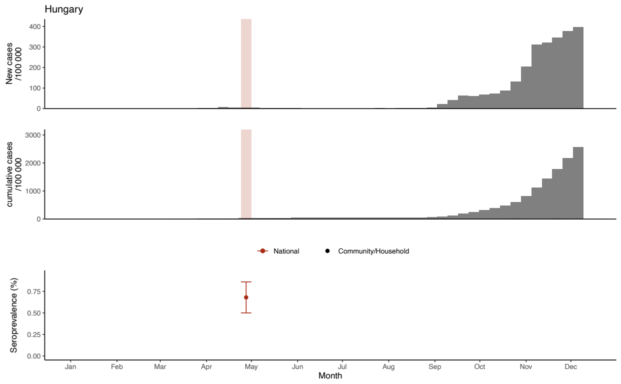

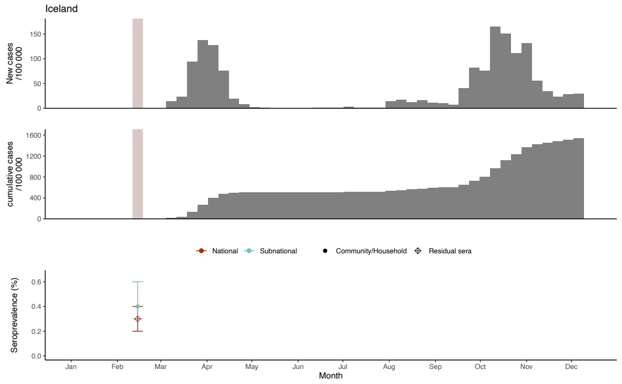

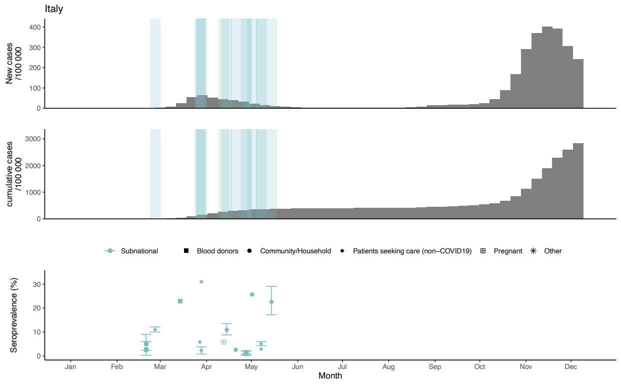

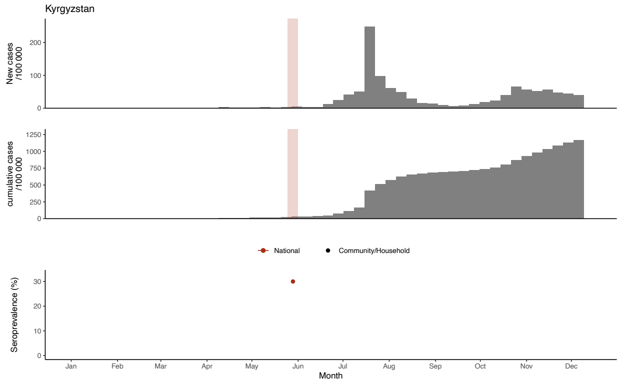

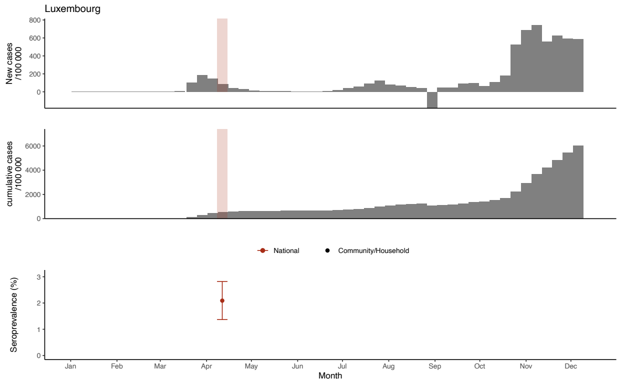

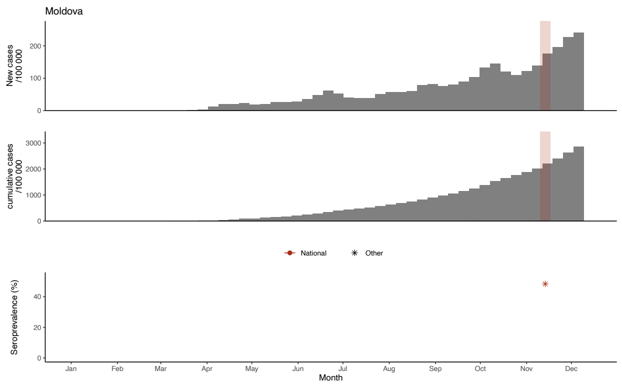

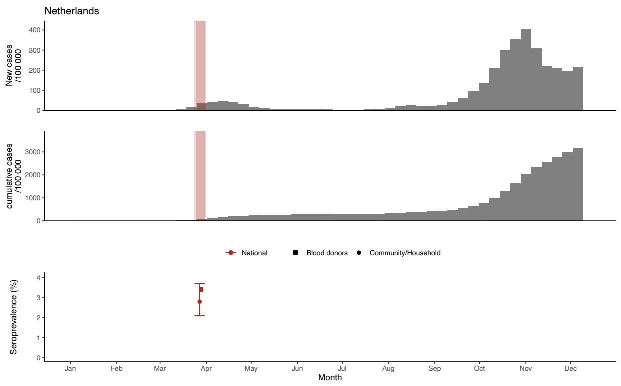

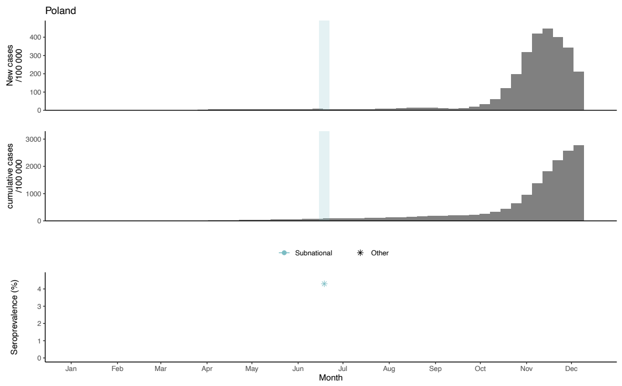

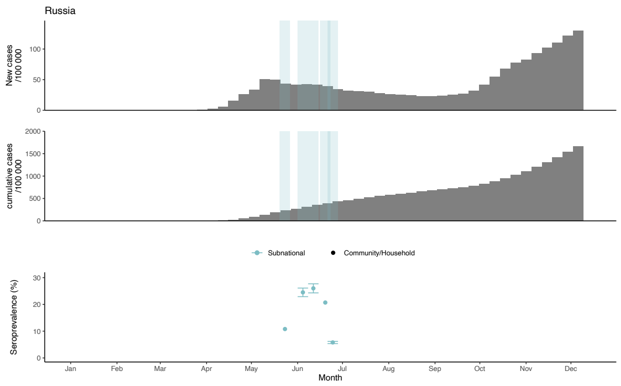

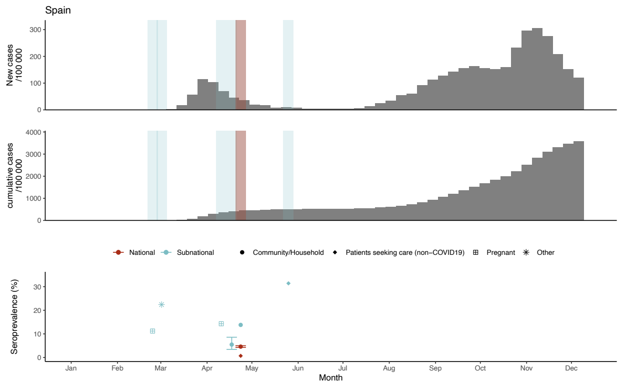

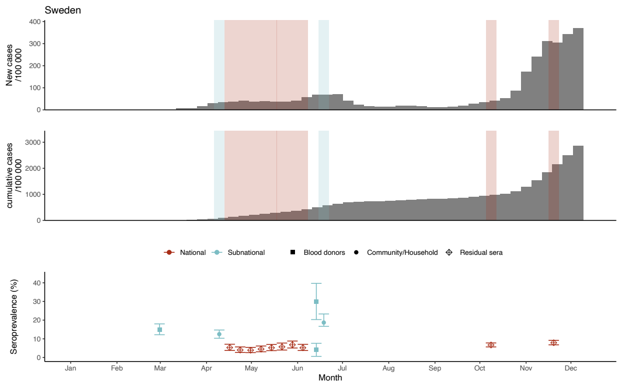

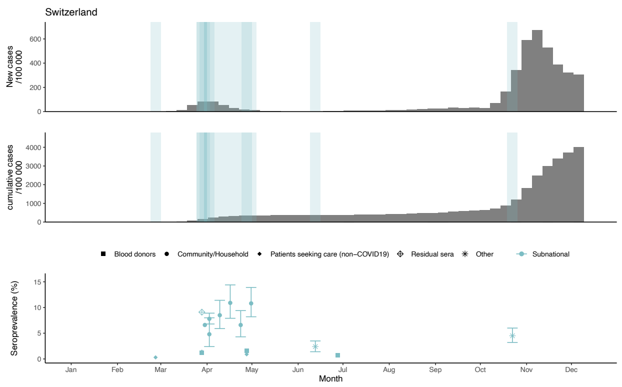

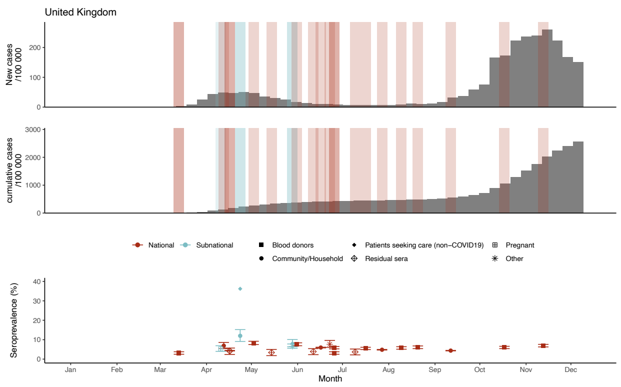
